# Pharmacovigilance and Mechanistic Analysis of Adverse Reaction Differences Across EGFR Tyrosine Kinase Inhibitor Generations in Non-Small Cell Lung Cancer

**DOI:** 10.64898/2026.10.16.26364955

**Authors:** Oluwadamilola Arojo, Alan M. Jones

## Abstract

**Background:** Epidermal Growth Factor Receptor (EGFR) tyrosine kinase inhibitors (TKIs) are widely used for EGFR-mutated non-small cell lung cancer (NSCLC) but have distinct toxicity profiles across generations.

**Aims:** To investigate relationships between EGFR-TKI physicochemical, pharmacokinetic and pharmacological properties and drug-specific adverse drug reactions (ADRs) using national and global pharmacovigilance data.

**Methods:** ADRs for erlotinib, afatinib and osimertinib were extracted from the UK MHRA Yellow Card scheme and WHO VigiAccess. Disproportionality analysis used reporting odds ratios (RORs). Signals were defined as ROR ≥2, lower 95% confidence interval >1 and ≥3 cases. Drug properties were curated from ChEMBL, emc, SmPC and EMA sources.

**Results:** VigiAccess contained 88,662 individual case safety reports (163,619 ADRs) and Yellow Card contained 752 reports (1,814 ADRs). Four System Organ Class (SOC) signals were identified in VigiAccess and three in Yellow Card, with five additional Yellow Card Preferred Term signals. Eye disorders showed the strongest consistent association with erlotinib (VigiAccess ROR 4.33; Yellow Card ROR 4.18). Osimertinib was associated with pneumonitis (ROR 7.67) and pulmonary embolism (ROR 3.94). Afatinib showed psychiatric disorder signals in both databases (ROR 3.2– 3.3) and diarrhoea (ROR 2.79). Neoplasm-related signals likely reflected confounding by indication.

**Conclusions:** EGFR-TKIs demonstrate generation-specific pharmacovigilance profiles. Integration with drug properties supports mechanistic plausibility for several signals, while the atypical afatinib psychiatric association warrants further investigation. Findings are hypothesis-generating and require confirmation in controlled observational or mechanistic studies.

**What is already known about this subject?:**

- First-, second-and third-generation EGFR-TKIs (erlotinib, afatinib and osimertinib) are established treatments for EGFR-mutated NSCLC but are associated with distinct adverse drug reaction (ADR) profiles that differ across generations.
- Known class-effect toxicities include dermatological, gastrointestinal, and ocular ADRs, attributed to EGFR inhibition in non-tumour epithelial tissues.
- Prior pharmacovigilance studies using the US FDA Adverse Event Reporting System (FAERS) have characterised EGFR-TKI safety signals, but UK-specific Yellow Card data have not been systematically analysed across all three generations, and the pharmacological basis for generation-specific differences remains incompletely understood.

**What this study adds:**

- Dual-database disproportionality analysis of MHRA Yellow Card and WHO VigiAccess data, using a pre-specified signal threshold (ROR ≥ 2, lower 95% CI > 1, n ≥ 3), confirmed generation-specific signals: eye disorders for erlotinib; psychiatric disorders and diarrhoea for afatinib; and pneumonitis and pulmonary embolism for osimertinib.
- Integration of target potency (pIC50), ErbB family selectivity, blood brain barrier (BBB) penetration and CYP_450_ interaction data with the pharmacovigilance signals provides mechanistic plausibility, linking them to variations in binding reversibility, wild-type EGFR inhibition, and off-target kinase activity.
- Applying a pre-specified, conservative signal threshold distinguished genuine disproportionate signals from elevated but non-significant reporting and identified neoplasm-related findings as confounded by indication rather than true drug safety signals highlighting the importance of rigorous signal definition in oncology pharmacovigilance.

## Introduction

Lung cancer remains a leading cause of cancer-related deaths worldwide, accounting for 18% of cancer deaths in 2020 [1]. It arises from uncontrolled proliferation of abnormal epithelial cells within pulmonary tissue, leading to malignant tumour formation [1]. Non-small cell lung cancer (NSCLC) is the most common subtype, accounting for 85% of all lung cancer diagnoses [2]. When detected at an early, localised stage, the five-year survival rate for NSCLC is approximately 65% [3]; however, many patients present at an advanced or metastatic stage, reducing five-year survival to 9%, with 30% of patients dying within three months of diagnosis [4,5].

Activating mutations in the epidermal growth factor receptor (EGFR) gene occur in 10–15% of NSCLC cases in Western populations and in up to 50% in Asian populations and are associated with poor clinical prognosis [2]. These mutations drive continuous activation of the EGFR tyrosine kinase, promoting tumour growth, angiogenesis, and metastasis through persistent activation of pro-oncogenic signalling pathways, including RAS-RAF-MEK-ERK/MAPK and AKT-PI3K-mTOR, via receptor overexpression, amplification, and uncontrolled downstream signalling [6–8].

Targeted therapy with EGFR tyrosine kinase inhibitors (TKIs) has been developed to selectively disrupt these tumour-promoting signals [9]. EGFR-TKIs inhibit receptor signalling by binding to the adenosine triphosphate (ATP)-binding site of the EGFR kinase domain, disrupting downstream signal transmission [9]. Three generations of EGFR-TKI are in clinical use. This study examines one representative agent *per* generation: erlotinib (first generation), afatinib (second generation) and osimertinib (third generation), licensed in the UK in 2005, 2013 and 2021 respectively [10–13]. All three are indicated for EGFR-mutated NSCLC, erlotinib is also licensed for pancreatic cancer [14–16]. First-generation EGFR-TKIs reversibly inhibit EGFR at the ATP-binding site; second-generation agents irreversibly inhibit EGFR and other ErbB family members to overcome resistance mechanisms [9,17]; and third-generation agents are designed to irreversibly and selectively target mutant EGFR, including the *T790M* resistance mutation, while sparing wild-type (WT) EGFR, thereby reducing off-target toxicity [17].

The World Health Organization (WHO) defines an adverse drug reaction (ADR) as “a response to a drug which is noxious and unintended, and which occurs at doses normally used for prophylaxis, diagnosis or therapy of disease, or for modification of physiological function” [18]. ADRs account for 7% of UK hospital admissions [19], and patients receiving chemotherapy experience at least one ADR during treatment [20]; approximately 16% of ADR-related hospital admissions are associated with antineoplastic drugs [21]. ADRs may necessitate dose reduction or treatment discontinuation, potentially compromising efficacy, and outcomes [22]. Although EGFR-TKIs have significantly improved survival in EGFR-mutated NSCLC, they are associated with a range of ADRs, including severe toxicities [23].

Despite sharing a common mechanism of action, first-, second-and third-generation EGFR-TKIs exhibit distinct ADR profiles, the mechanistic basis for which is incompletely understood but may relate to differences in target selectivity, off-target activity, chemical structure, and pharmacokinetic behaviour. Recent pharmacovigilance studies have begun to characterise EGFR-TKI safety profiles using large-scale spontaneous reporting databases. Huang et al. conducted a disproportionality analysis of four EGFR-TKIs using the US FDA Adverse Event Reporting System (FAERS), identifying generation-specific signals across 27,123 reports [24]. More recently, Shi et al. combined a network meta-analysis of 46 randomised controlled trials with a FAERS disproportionality analysis covering 75,196 patients and over 200,000 adverse event occurrences across six EGFR-TKIs [25]. Both studies provided valuable insight into the comparative safety of EGFR-TKIs; however, they relied exclusively on FAERS, which may not fully reflect prescribing patterns, reporting practices or patient populations in other healthcare systems. Furthermore, neither study systematically integrated the pharmacovigilance signals with the underlying pharmacological, physicochemical, and pharmacokinetic properties of the drugs to explore mechanistic explanations for the observed differences.

*This study* therefore aimed to investigate ADR patterns associated with EGFR-TKIs through analysis of both national (UK) and global pharmacovigilance data, and to explore whether pharmacological and molecular factors contribute to generation-specific differences in safety profiles. A dual-database approach was employed, using the MHRA Yellow Card scheme (which captures UK-specific individual case safety reports not represented in FAERS) alongside the WHO VigiAccess database (which aggregates worldwide reports), enabling triangulation of signals across independent data sources. Disproportionality analysis was integrated with comparative profiling of target potency (IC_50_ against EGFR variants and ErbB family kinases), binding reversibility, blood–brain barrier (BBB) penetration, CYP_450_ interaction profiles and key physicochemical properties, providing a mechanistic framework for interpreting the observed ADR patterns. We hypothesised that differences in ADR profiles across EGFR-TKI generations are driven by variations in target selectivity, pharmacokinetics, and molecular structure.

## Methods

### Pharmacovigilance data sources and ADR extraction

ADR data were retrieved from two public pharmacovigilance databases: (i) the national MHRA Yellow Card scheme [26], representing UK individual case safety reports (ICSRs); and (ii) the WHO VigiAccess database [27], representing worldwide ICSRs collated by the Uppsala Monitoring Centre. Both databases collect spontaneous reports of suspected ADRs for medicines, vaccines and medical devices submitted by healthcare professionals, patients, carers, and pharmaceutical companies. In both databases, ADRs are coded according to the Medical Dictionary for Regulatory Activities (MedDRA) hierarchy, version 28.1 [28]. Analyses were conducted at System Organ Class (SOC) level for both databases, with additional Preferred Term (PT)-level analysis performed for the MHRA dataset. WHO VigiAccess data were restricted to SOC-level analysis as the public interface does not permit PT-level case-level extraction.

Data were extracted manually into Microsoft Excel (version 16.104) spreadsheets. Separate datasets were created for each database, in which all drug-event pairs were recorded, including drug name, SOC (and PT for MHRA data) and the case counts required to construct ROR contingency tables. The dataset included all suspected ADRs associated with the three EGFR-TKIs from the year of UK licensing of each drug (erlotinib 2005, afatinib 2013, osimertinib 2021) until October 2025. All ADRs were analysed under the assumption that the drugs were prescribed for the licensed primary indication of NSCLC; however, erlotinib is also licensed for pancreatic cancer in the UK, and neither database permits confirmation of indication for individual reports (*see Limitations*). One representative drug *per generation* was selected *a priori* to enable a focused mechanistic comparison rather than an exhaustive survey of all licensed EGFR-TKIs.

### Inclusion and exclusion criteria

Inclusion and exclusion criteria (**Table 1**) were applied to minimise the inclusion of non-clinical, non-quantifiable or non-drug-related data. Because EGFR-TKIs are used exclusively in patients with active malignancy, SOC terms relating to disease progression (“Neoplasms benign, malignant and unspecified [incl. cysts and polyps]”) and its associated Preferred Terms (e.g. “metastases,” “malignant neoplasm progression”) are highly susceptible to confounding by indication, because disease progression and drug-related harm cannot be distinguished in spontaneous report data. These terms were therefore retained in the descriptive dataset but are explicitly labelled as confounded in the Results and Discussion and are not interpreted as drug safety signals.

**Table 1.** Inclusion and exclusion criteria applied to pharmacovigilance ADR data.

| Inclusion | Exclusion |
| --- | --- |
| Quantifiable numerical case counts | Administration site issues |
| Reports mapped to a recognised SOC and PT | Product use errors / procedural errors |
| The ADR must also occur with the other two drugs (to enable comparison) | Investigations (laboratory/diagnostic findings without a defined ADR) |
| The ADR must occur at least three times for the drug | Surgical and medical procedures |
| The database must contain background (denominator) counts | Neoplasm progression terms (retained for transparency but flagged as confounded by indication and not used as safety signals) |

### Disproportionality analysis

Owing to the nature of the available data, disproportionality analysis (DA) was used to identify statistically disproportionate ADR reporting; DA is a well-established, hypothesis-generating tool that requires confirmation through more rigorous epidemiological or mechanistic studies [30]. The reporting odds ratio (ROR) was selected as it is a widely used and validated disproportionality measure in pharmacovigilance [30,31], calculated from a 2×2 contingency table (**Tables 2-3**) comparing reports of a given ADR for the drug of interest against all other ADR reports for that drug, and against reports of the same ADR and all other ADRs for the comparator drugs.

**Table 2.** Two-by-two contingency table used for ROR calculation.

|  | Reports with ADR of interest | All other ADR reports |
| --- | --- | --- |
| Drug of interest | a | b |
| All other drugs | c | d |

**Table 3.** Algorithms used for signal detection.

| Statistic | Formula | Criterion |
| --- | --- | --- |
| ROR | $(a + 0.5) \times (d + 0.5) / [(b + 0.5) \times (c + 0.5)]$ | see below |
| Approx. SE | $\sqrt{1/(a+0.5) + 1/(b+0.5) + 1/(c+0.5) + 1/(d+0.5)}$ | — |
| Ln(ROR) | ln(ROR) | — |
| 95% CI lower | $\exp[\ln(\text{ROR}) - 1.96 \times \text{Approx. SE}]$ | — |
| 95% CI upper | $\exp[\ln(\text{ROR}) + 1.96 \times \text{Approx. SE}]$ | — |
| Signal? | IF (AND(95% CI lower > 1, a ≥ 3, ROR ≥ 2), “Signal detected”, “No signal”) | ROR ≥ 2, lower 95% CI > 1 and n ≥ 3 |

Notes: ROR, reporting odds ratio; Approx. SE, approximate standard error; CI, confidence interval. “a” = number of reports of the ADR of interest for drug X; “b” = number of other ADR reports for drug X; “c” = number of reports of the ADR of interest for all other drugs; “d” = number of all other ADR reports for all other drugs.

A formal safety signal was defined a priori using the standard criteria of Huang et al. [24]: ROR ≥ 2, lower limit of the 95% CI > 1, and n ≥ 3 cases. Associations with a lower 95% CI > 1 but ROR between 1 and 2 were considered elevated reporting of potential interest but did not meet the pre-specified signal threshold and are reported separately as such, rather than as confirmed signals; this distinction was applied consistently throughout the Results and Discussion. No correction for multiple comparisons was applied, consistent with the hypothesis-generating, exploratory nature of disproportionality analysis [30,31]; this is acknowledged as a limitation.

Forest plots were generated in *R*Studio (2023) using the ggplot2 package to visualise signal magnitude and precision. As the focus of this exploratory study was the comparison of ADR patterns between the three EGFR-TKIs, adjustment for other confounding variables (e.g., age, sex, treatment line, concomitant medication) was not possible with the aggregated data available and is discussed as a limitation.

### Sensitivity analysis: defined daily dose (DDD)-adjusted reporting

Because the three EGFR-TKIs were licensed in different years and have different prescribing volumes, raw case counts are influenced by drug utilisation as well as by true differences in toxicity. As a sensitivity analysis, ADR reporting rates were additionally expressed per 100,000 defined daily doses (DDDs) dispensed, using the WHO Collaborating Centre for Drug Statistics Methodology Anatomical Therapeutic Chemical (ATC)/DDD classification system [29] to standardise for exposure. Chi-squared (χ²) tests were used to compare standardised ADR-per-100,000-DDD rates between drug pairs, with a p-value <0.001 used as the threshold for statistical significance (**Supplementary Information A1-A4**). This analysis focused on whether reporting rates differed between drugs overall, rather than on signal detection for individual ADRs; because of the exploratory nature of this sensitivity analysis, the limited availability of prescribing denominator data, the lack of adjustment for potential confounders, and the relatively low case counts for some ADRs, no correction for multiple comparisons was applied to the chi-squared analysis, and results should be interpreted as supportive rather than confirmatory of the ROR-based disproportionality findings.

### Target potency, physicochemical and pharmacokinetic data

Target potency (IC_50_) data for erlotinib, afatinib and osimertinib against wild-type EGFR, clinically relevant EGFR mutations and other ErbB family kinases were extracted from ChEMBL [32]. Only numerical IC_50_ values from functional or cytotoxicity assays measured between 48 and 72 hours, with confirmed compound identity, were included; where multiple entries existed, the median IC50 was calculated to reduce the influence of outliers, and values were log-transformed (pIC_50_ = -log_10_[IC_50_ in M]) for comparative analysis. Where available, ChEMBL-derived potency values were cross-checked against independent *in vitro* mutant-specificity studies [33,34] to confirm concordance with the published literature.

Physicochemical properties, pharmacokinetic parameters and transporter data were obtained from the UK Summary of Product Characteristics (SmPC) via the electronic medicines compendium (emc) [14–16], EMA European Public Assessment Reports (EPARs) [10–12], and peer-reviewed pharmacokinetic literature [35,38]; where parameters were reported as ranges across studies, the median value is reported, with the original units and source indicated. Where a reported concentration required conversion from mass concentration (ng/mL) to molar units (nM) for direct comparison with IC_50_ potency values, the following conversion was applied:

*Concentration (nM) = [Concentration (ng/mL) / Molecular weight (g/mol)] × 1000*

Steady-state peak plasma concentration (C_max_) values for each drug were taken from the pharmacokinetics section of the relevant UK SmPC, using the value reported for the standard licensed NSCLC dose at steady state where available, in preference to single-dose or non-standard-dose values; where only a mass concentration was reported in the source document, this was converted to nM using the formula above, and the original reported value and units are also given in **Table 4** for transparency and independent verification.

**Table 4.**
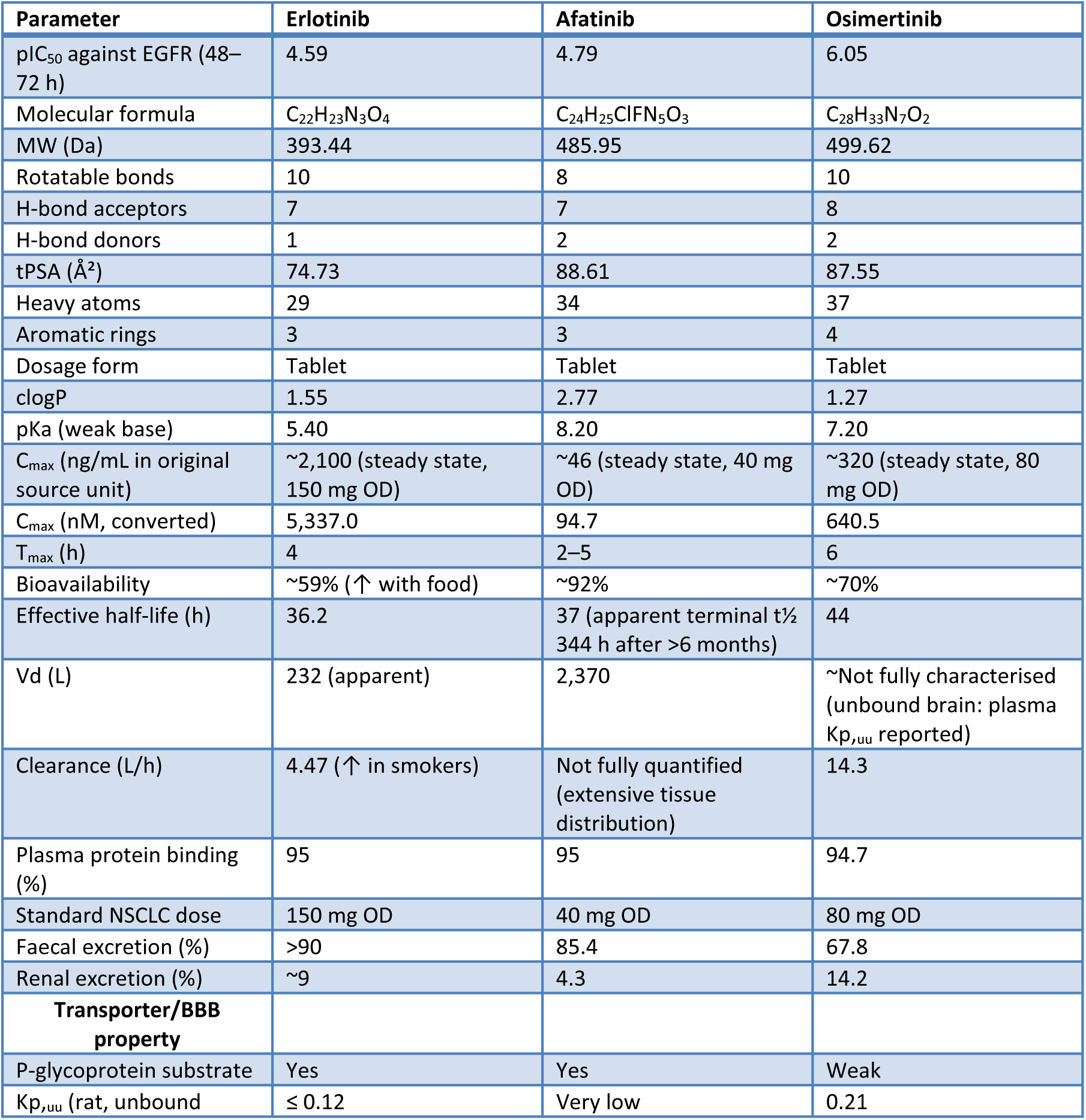
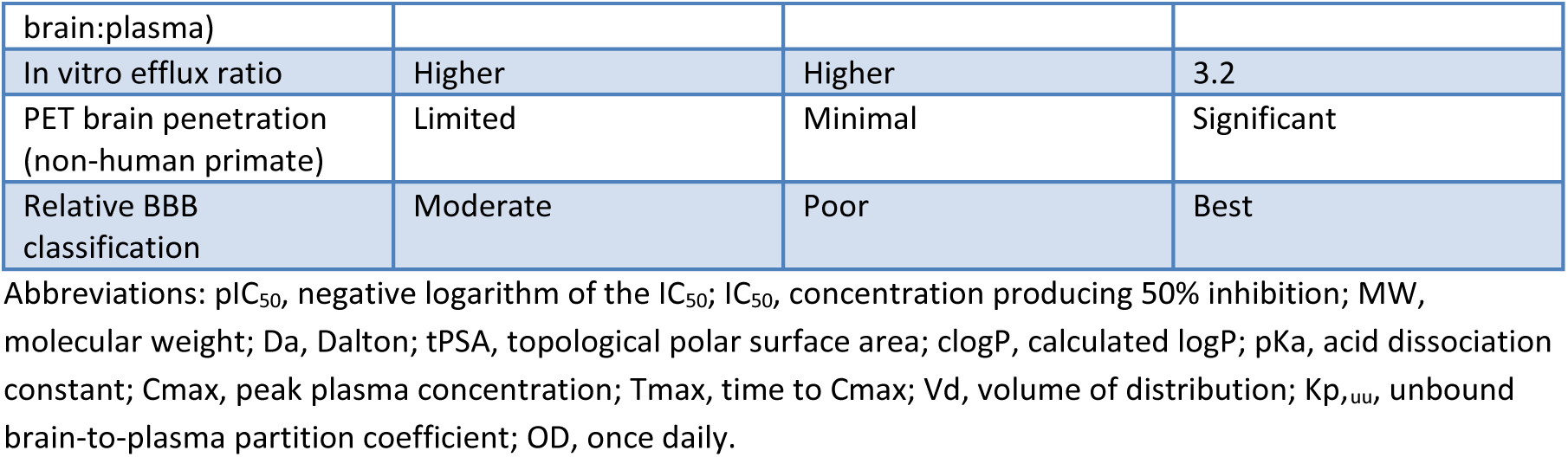
Physicochemical, blood–brain barrier, pharmacokinetic and pharmacological properties of the three EGFR-TKIs.

### 2.6 Ethics approval

This study used only publicly available, aggregated and anonymised pharmacovigilance data and did not involve identifiable patient information; ethical approval was therefore not required.

## Results

### Physicochemical, pharmacological, and pharmacokinetic properties

The physicochemical, pharmacological, and pharmacokinetic properties of erlotinib, afatinib and osimertinib are summarised in **Table 4**, and the shared and distinguishing chemical features are illustrated in **Figure 1**. The three drugs differed across all major property classes. Osimertinib exhibited the highest potency against mutant EGFR (pIC_50_ = 6.05) compared with erlotinib (pIC_50_ = 4.59) and afatinib (pIC_50_ = 4.79), whereas erlotinib showed the highest potency against wild-type (WT) EGFR (IC50 = 2 nM); osimertinib was designed to spare WT EGFR (IC_50_ = 938 nM). Afatinib was the most lipophilic agent (clogP 2.77) and had the highest topological polar surface area (tPSA 88.61 Å²), while erlotinib had the lowest molecular weight (393.44 Da) and tPSA (74.73 Å²).

**Figure 1.**
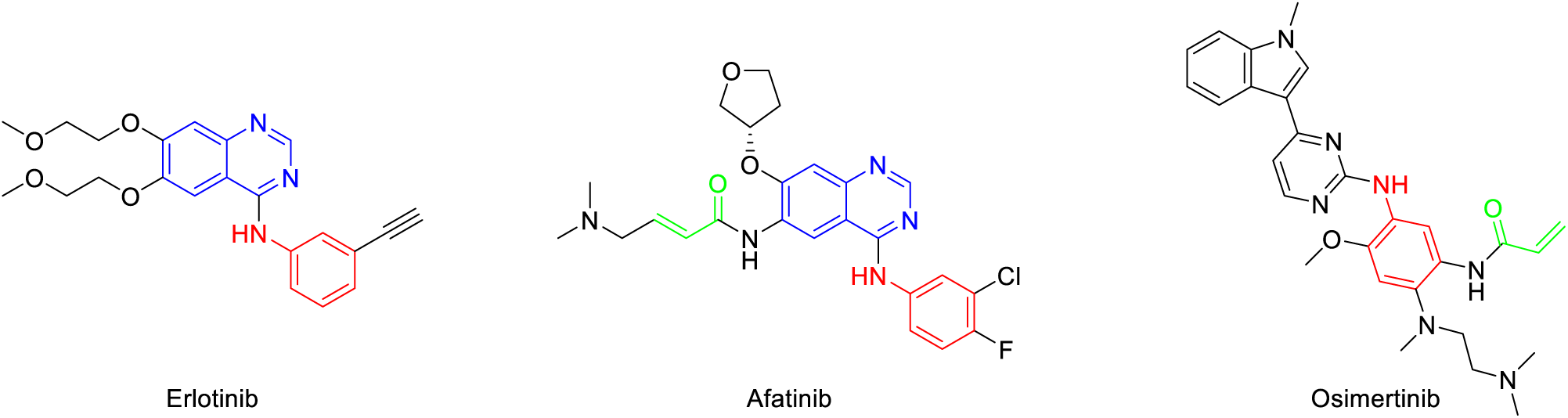
Chemical structures of the three EGFR-TKIs, highlighting the shared quinazoline core (erlotinib, afatinib; blue), the aniline moiety (red) and the Michael acceptor/acrylamide warhead unique to the covalent inhibitors afatinib and osimertinib (green).

Pharmacokinetic parameters also differed considerably (**Table 4**). At the standard licensed NSCLC dose at steady state, erlotinib achieved the highest peak plasma concentration (C_max_ ≈ 2.1 µg/mL, equivalent to approximately 5,337 nM in non-smoking patients at the 150 mg once-daily dose) [14], giving a Cmax:IC_50_ (WT EGFR) ratio of approximately 2,700-fold, by far the greatest of the three drugs, while afatinib had the lowest C_max_ (94.7 nM) but the largest volume of distribution (2,370 L) and highest apparent clearance; osimertinib had the longest effective half-life (44 h). All three drugs displayed high plasma protein binding (>94%). Afatinib’s elimination half-life requires clarification, the SmPC reports an effective half-life of approximately 37 h based on standard multiple-dose pharmacokinetic sampling, but a much longer apparent terminal half-life of 344 h (>14 days) was estimated in patients treated for more than 6 months, reflecting slow dissociation of afatinib from covalently bound EGFR/ErbB targets in tissues rather than a change in systemic clearance [15,36]. This distinction between effective (dosing-interval) and apparent terminal (tissue-release-limited) half-life is relevant to the sustained target engagement discussed later.

Transporter and BBB characteristics also differed across the three agents (**Table 4**, **Figure 1**). Erlotinib and afatinib were identified as substrates of P-glycoprotein and BCRP, with limited to minimal non-human primate PET brain penetration, whereas osimertinib showed weaker P-glycoprotein/BCRP interaction and greater BBB penetration, consistent with preclinical and clinical PET imaging data [38,52].

### Target affinity and selectivity

Target potency data for EGFR variants, ErbB family kinases and CYP450 enzymes are summarised in **Tables 5-7**. Osimertinib demonstrated the greatest potency against T790M-containing resistance mutations (IC_50_ = 2–11.44 nM), whereas erlotinib showed negligible activity against these variants (IC_50_ > 1,000 nM; **Table 5**). Afatinib was the most potent agent against common sensitising mutations (exon 19 deletion IC_50_ 0.36 nM; *L858R* IC_50_ 0.31 nM). All three drugs retained activity against the *G719C* variant, with osimertinib the most potent (IC_50_ = 0.002 nM). Erlotinib demonstrated the highest potency against WT EGFR (IC_50_ = 2 nM), consistent with the mechanistic interpretation.

**Table 5.** Comparative IC_50_ values (nM) of erlotinib, afatinib and osimertinib against wild-type EGFR and common activating/resistance mutations.

| Target | Erlotinib IC<sub>50</sub> (nM) | Afatinib IC<sub>50</sub> (nM) | Osimertinib IC<sub>50</sub> (nM) |
| --- | --- | --- | --- |
| EGFR Exon 19 del + T790M | >1000 | 141 | 2 |
| EGFR L858R (Exon 21) + T790M | >1000 | 196 | 11.44 |
| EGFR Exon 19 deletion | 23 | 0.36 | 12.92 |
| EGFR L858R (Exon 21) | 39 | 0.31 | 9 |
| EGFR wild type (ErbB1) | 2 | 31 | 938 |
| EGFR G719C | 0.03 | 0.40 | 0.002 |

**Table 6.** Selectivity profile of erlotinib, afatinib and osimertinib across ErbB family kinases (IC_50_, nM).

| Kinase | Erlotinib IC<sub>50</sub> (nM) | Afatinib IC<sub>50</sub> (nM) | Osimertinib IC<sub>50</sub> (nM) |
| --- | --- | --- | --- |
| ErbB2 (HER2) | 5.90 | 14 | 4.80 |
| ErbB4 | 7.60 | 2 | 46 |
| ErbB3 | 10.00 | No significant activity | 2.10 |

**Table 7.** CYP_450_ interaction profiles of erlotinib, afatinib and osimertinib.

| Enzyme | Erlotinib | Afatinib | Osimertinib |
| --- | --- | --- | --- |
| CYP3A4 | Substrate | No significant interaction | Substrate |
| CYP3A5 | Substrate | No significant interaction | Substrate |
| CYP1A2 | Substrate (induced by smoking) | No significant interaction | No significant interaction |
| CYP2C8 | Substrate | No significant interaction | No significant interaction |
| CYP1B1 | Substrate | No significant interaction | No significant interaction |

Inhibition of other ErbB family kinases varied across the three drugs (**Table 6**). Afatinib showed the broadest ErbB activity, potently inhibiting ErbB2/HER2 and ErbB4, but did not meaningfully inhibit ErbB3. Erlotinib inhibited all four ErbB family members, including HER2 (IC_50_ = 5.90 nM). Osimertinib showed potent ErbB3 activity (IC_50_ = 2.10 nM) but weaker ErbB4 inhibition (IC_50_ = 46 nM) than afatinib (IC_50_ = 2 nM) and retained inhibitory activity against HER2 (IC_50_ = 4.80 nM).

Erlotinib had the most extensive CYP_450_ interaction profile, acting as a substrate of CYP3A4, CYP3A5, CYP1A2, CYP2C8 and CYP1B1 (**Table 7**); its CYP1A2-mediated metabolism is of clinical relevance because smoking induces CYP1A2 and reduces erlotinib exposure. Osimertinib was a substrate of CYP3A4 and CYP3A5 only, generating two active circulating metabolites (AZ5104 and AZ7550) with retained EGFR activity (**Figure 2**). Afatinib showed no clinically significant CYP-mediated metabolism, being eliminated predominantly unchanged via biliary/faecal excretion (85.4% of an oral dose recovered in faeces, with the parent compound accounting for 88% of the recovered dose) [15].

**Figure 2.**
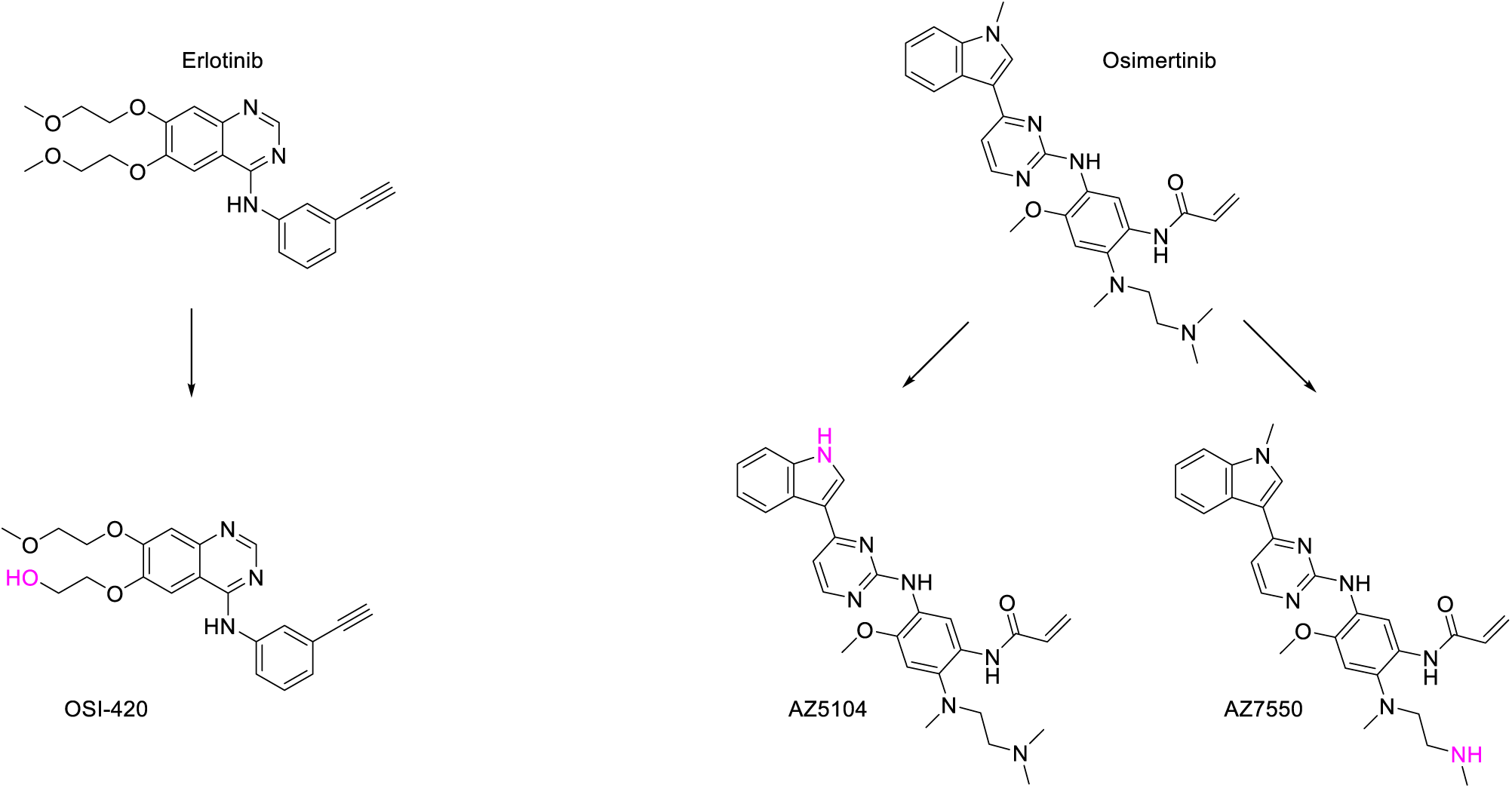
Major circulating metabolites of erlotinib and osimertinib, generated primarily via CYP3A4/CYP3A5-mediated oxidation.

### Disproportionality analysis

WHO VigiAccess contained *n* = 88,662 ICSRs, comprising *n* = 163,619 suspected ADRs associated with erlotinib, afatinib and osimertinib. VigiAccess does not provide stratification by seriousness or fatality. Between date of license in the UK and October 2025, the MHRA Yellow Card scheme contained *n* = 752 ICSRs associated with the three drugs, comprising *n* = 1,814 suspected ADRs (erlotinib *n* = 940; afatinib *n* = 295; osimertinib *n* = 579). Of the Yellow Card reports, 87.5% were classified as serious, 17.7% were fatal, and 98.9% were submitted by healthcare professionals.

### Signal detection at SOC level

Applying the pre-specified signal criteria (ROR ≥ 2, lower 95% CI > 1, n ≥ 3; Section 2.3), four SOC-level signals were confirmed in VigiAccess and three in the Yellow Card scheme (**Tables 8-9** and **Figures 3-4**). A number of additional SOC-level associations showed a lower 95% CI > 1 but an ROR between 1 and 2; these are reported in **Tables 8-9** for transparency and comparison across databases but are classified as elevated reporting rather than confirmed signals, consistent with the pre-specified threshold.

**Figure 3:**
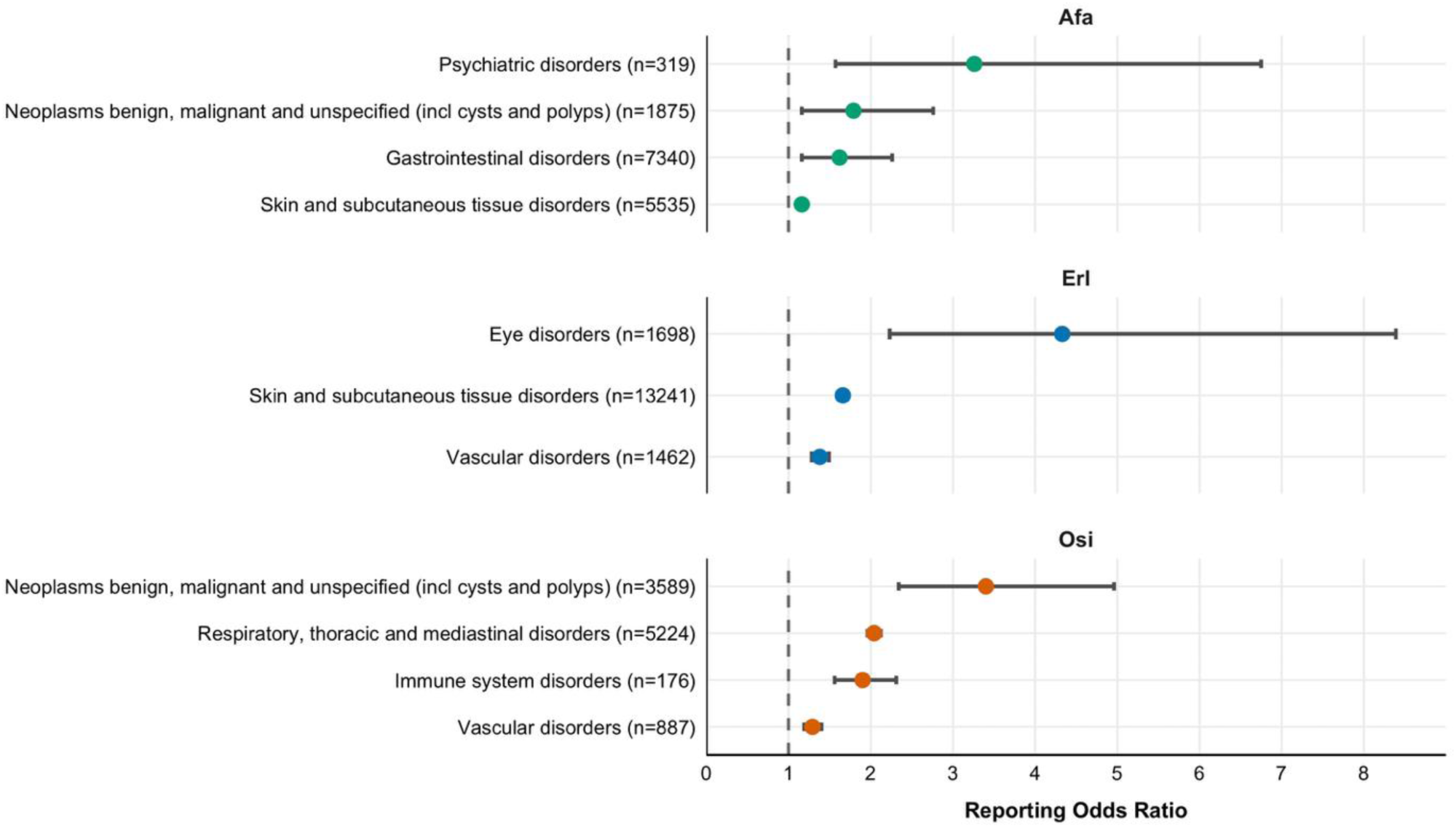
Forest plot of RORs for SOC-level associations identified in the WHO VigiAccess database. Erl, erlotinib; Afa, afatinib; Osi, osimertinib; n, number of events.

**Figure 4.**
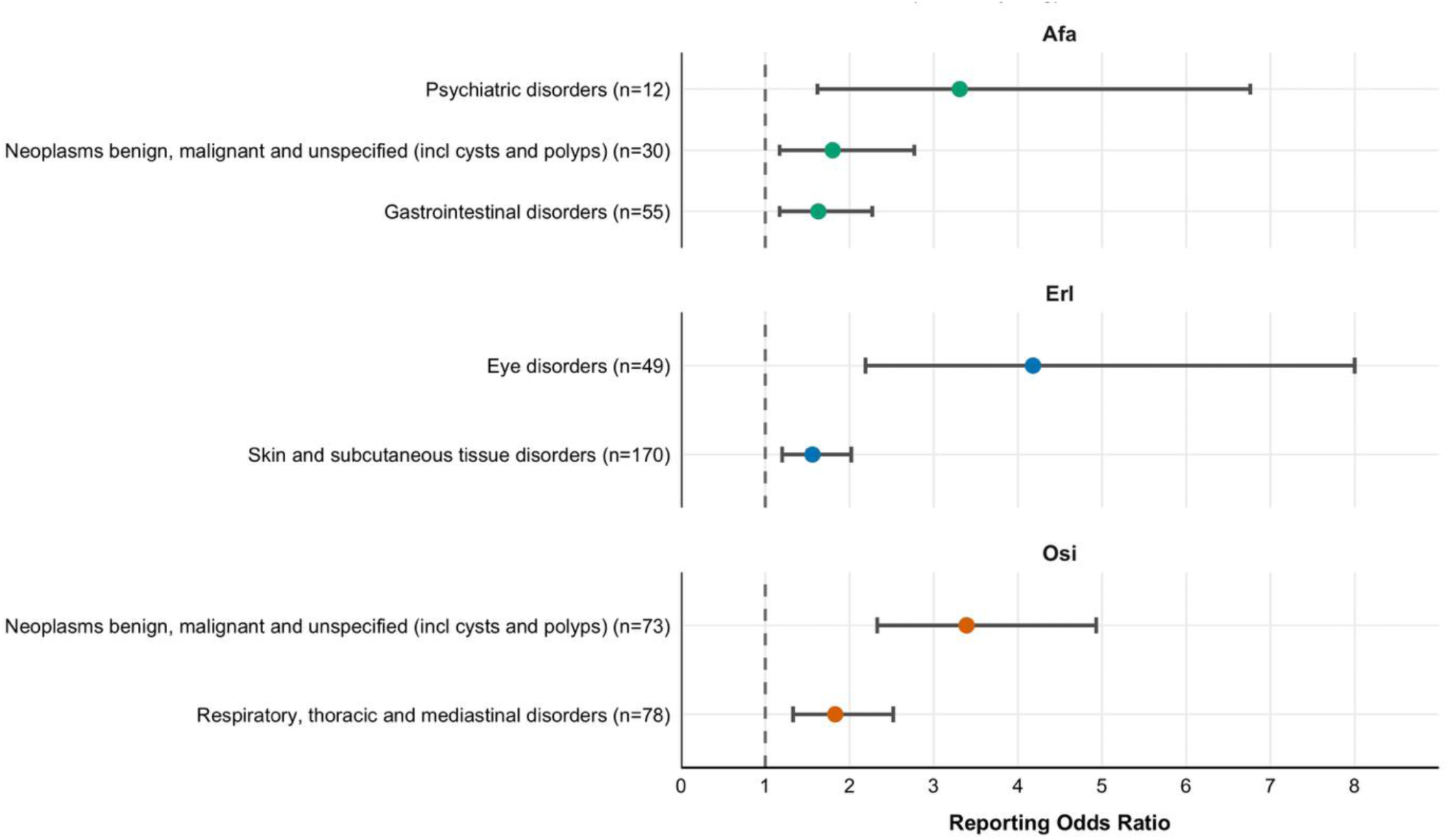
Forest plot of RORs for SOC-and PT-level associations identified in the MHRA Yellow Card scheme. Erl, erlotinib; Afa, afatinib; Osi, osimertinib; n, number of events.

**Table 8.** WHO VigiAccess disproportionality results at System Organ Class (SOC) level.

| Drug | SOC | n | ROR | ROR 95% CI | Signal (ROR≥2, lower CI$>$1, n≥3) |
| --- | --- | --- | --- | --- | --- |
| Erlotinib | Eye disorders | 1698 | 4.33 | 2.23–8.39 | Yes |
| Afatinib | Gastrointestinal disorders | 7340 | 1.62 | 1.16–2.26 | No (elevated only) |
| Osimertinib | Neoplasms benign, malignant, and unspecified | 3589 | 3.40 | 2.34–4.96 | Statistically yes – confounded by indication |
| Afatinib | Neoplasms benign, malignant, and unspecified | 1875 | 1.79 | 1.16–2.76 | No (elevated only; also confounded) |
| Afatinib | Psychiatric disorders | 319 | 3.26 | 1.57–6.75 | Yes |
| Osimertinib | Respiratory, thoracic, and mediastinal disorders | 5224 | 2.04 | 1.96–2.12 | Yes |
| Afatinib | Skin and subcutaneous tissue disorders | 5535 | 1.16 | 1.12–1.20 | No (elevated only) |
| Erlotinib | Skin and subcutaneous tissue disorders | 13241 | 1.66 | 1.61–1.71 | No (elevated only) |
| Osimertinib | Vascular disorders | 887 | 1.29 | 1.19–1.40 | No (elevated only) |
| Erlotinib | Vascular disorders | 1462 | 1.38 | 1.28–1.49 | No (elevated only) |
| Osimertinib | Immune system disorders | 176 | 1.90 | 1.56–2.31 | No (elevated only) |

**Table 9.** MHRA Yellow Card scheme disproportionality results at SOC and Preferred Term (PT) level.

| Drug | SOC / PT | n | ROR | ROR 95% CI | Signal |
| --- | --- | --- | --- | --- | --- |
| Erlotinib | Eye disorders (SOC) | 49 | 4.18 | 2.19–8.00 | Yes |
| Erlotinib | Lid, lash and lacrimal structural disorders (PT) | 12 | 7.84 | 1.44–42.78 | Yes (wide CI, small n) |
| Afatinib | Gastrointestinal disorders (SOC) | 55 | 1.63 | 1.17–2.27 | No (elevated only) |
| Afatinib | Diarrhoea (PT) | 28 | 2.79 | 1.74–4.46 | Yes |
| Osimertinib | Neoplasms benign, malignant, and unspecified (SOC) | 73 | 3.39 | 2.33–4.93 | Statistically yes – confounded by indication |
| Osimertinib | Metastases (PT) | 27 | 3.07 | 1.70–5.54 | Statistically yes – confounded |
| Osimertinib | Malignant neoplasm progression (PT) | 38 | 4.64 | 2.64–8.16 | Statistically yes – confounded |
| Afatinib | Neoplasms benign, malignant, and unspecified (SOC) | 30 | 1.80 | 1.17–2.77 | No (elevated only; confounded) |
| Afatinib | Malignant neoplasm progression (PT) | 15 | 2.02 | 1.11–3.67 | Statistically yes – confounded |
| Afatinib | Psychiatric disorders (SOC) | 12 | 3.31 | 1.62–6.76 | Yes |
| Osimertinib | Respiratory, thoracic, and mediastinal disorders (SOC) | 78 | 1.83 | 1.33–2.52 | No (elevated only at SOC) |
| Osimertinib | Pneumonitis (PT) | 29 | 7.67 | 3.55–16.57 | Yes |
| Osimertinib | Pulmonary embolism (PT) | 6 | 3.94 | 1.07–14.52 | Yes (wide CI, small n) |
| Erlotinib | Skin and subcutaneous tissue disorders (SOC) | 170 | 1.56 | 1.20–2.02 | No (elevated only) |
| Erlotinib | Rash (PT) | 77 | 1.81 | 1.22–2.67 | No (elevated only) |

Notes (Tables 8–9): Signal defined as ROR $\geq 2$ , $n \geq 3$ and lower 95% CI $> 1$ . Entries meeting the statistical criterion but confounded by indication (neoplasm-related terms) are labelled accordingly and are not interpreted as drug safety signals (Section 4.2.1). CI, confidence interval; PT, Preferred Term; ROR, reporting odds ratio; SOC, System Organ Class.

### Confirmed SOC-level signals were as follows

#### Erlotinib

Eye disorders: the strongest signal detected for any drug–SOC combination in this study, confirmed in both databases (VigiAccess ROR 4.33, 95% CI 2.23–8.39, *n* = 1,698; Yellow Card ROR 4.18, 95% CI 2.19–8.00, *n* = 49).

#### Afatinib

Psychiatric disorders: confirmed in both databases (VigiAccess ROR 3.26, 95% CI 1.57–6.75, *n* = 319; Yellow Card ROR 3.31, 95% CI 1.62–6.76, *n* = 12).

#### Osimertinib

Respiratory, thoracic and mediastinal disorders: confirmed in VigiAccess (ROR 2.04, 95% CI 1.96–2.12, *n* = 5,224); the equivalent Yellow Card association (ROR 1.83, 95% CI 1.33–2.52, *n* = 78) had a lower 95% CI > 1 but did not meet the ROR ≥ 2 threshold and is classified as elevated reporting at SOC level (the corresponding PT-level analysis, did meet the signal threshold).

Associations with a lower 95% CI > 1 but ROR < 2, and therefore classified as elevated reporting rather than confirmed signals, included: gastrointestinal disorders for afatinib (VigiAccess ROR 1.62, 95% CI 1.16–2.26; Yellow Card ROR 1.63, 95% CI 1.17–2.27); skin and subcutaneous tissue disorders for erlotinib (VigiAccess ROR 1.66, 95% CI 1.61–1.71; Yellow Card ROR 1.56, 95% CI 1.20–2.02) and afatinib (VigiAccess ROR 1.16, 95% CI 1.12–1.20); vascular disorders for erlotinib (ROR 1.38, 95% CI 1.28–1.49) and osimertinib (ROR 1.29, 95% CI 1.19–1.40) in VigiAccess; and immune system disorders for osimertinib in VigiAccess (ROR 1.90, 95% CI 1.56–2.31).

Neoplasms benign, malignant and unspecified (incl. cysts and polyps) showed elevated reporting for osimertinib (VigiAccess ROR 3.40, 95% CI 2.34–4.96; Yellow Card ROR 3.39, 95% CI 2.33– 4.93) and afatinib (VigiAccess ROR 1.79; Yellow Card ROR 1.80), formally meeting the ROR ≥ 2 threshold for osimertinib. However, as pre-specified, this association is interpreted as confounding by indication rather than a genuine drug safety signal, because EGFR-TKIs are not established to cause secondary malignancy, and osimertinib is frequently used in later-line, *T790M*-resistant disease where progression is more common. Forest plots of all SOC-level associations are shown in **Figure 3** (VigiAccess) and **Figure 4** (Yellow Card scheme).

### Signal detection at PT level (MHRA Yellow Card)

MHRA Yellow Card data were further analysed at PT level (**Table 9**). Excluding neoplasm-related terms (confounded by indication), five additional PT-level signals meeting the pre-specified criteria were identified: lid, lash and lacrimal structural disorders for erlotinib (ROR 7.84, 95% CI 1.44–42.78, *n* = 12); diarrhoea for afatinib (ROR 2.79, 95% CI 1.74–4.46, *n* = 28); and pneumonitis (ROR 7.67, 95% CI 3.55–16.57, *n* = 29) and pulmonary embolism (ROR 3.94, 95% CI 1.07–14.52, *n* = 6) for osimertinib. Rash for erlotinib (ROR 1.81, 95% CI 1.22–2.67, *n* = 77) showed elevated reporting but did not meet the ROR ≥ 2 threshold. The confidence intervals for several signals were wide, particularly those with small case counts (lid/lash/lacrimal disorders, *n* = 12; pulmonary embolism, *n* = 6), and these findings should be interpreted with appropriate caution. Forest plots of PT-level signals are presented in **Figure 4**.

Overall, eye, skin, gastrointestinal and vascular associations were consistently directionally elevated across both databases, supporting reasonable external consistency of the dataset, even where the magnitude of the association did not meet the pre-specified ROR ≥ 2 signal threshold in one or both databases.

## Discussion

This pharmacovigilance study identified both expected and atypical ADR signals associated with first-, second-and third-generation EGFR-TKIs in NSCLC. Using a pre-specified, conservative signal threshold (ROR ≥ 2, lower 95% CI > 1, n ≥ 3), ocular toxicity for erlotinib, psychiatric disorders for afatinib, and pneumonitis and pulmonary embolism for osimertinib were confirmed as signals in at least one database, with eye disorders (erlotinib) and psychiatric disorders (afatinib) confirmed in both. Gastrointestinal and dermatological associations, long recognised as class-effect toxicities of EGFR-TKIs and documented in the British National Formulary [39–41], were consistently elevated across both databases but did not meet the pre-specified ROR ≥ 2 threshold at SOC level, with the exception of afatinib-associated diarrhoea at PT level.

These findings are broadly consistent with previous FAERS-based analyses. Huang et al. identified generation-specific signals across four EGFR-TKIs, including dermatological toxicity for erlotinib and gastrointestinal toxicity for afatinib [24]. Shi et al. confirmed these patterns across a larger dataset of over 200,000 adverse event occurrences and additionally identified cardiac and hepatic signals for osimertinib [25]. The present study extends these findings in three respects: first, by providing UK-specific Yellow Card data reflecting national prescribing and reporting practices not captured by FAERS; second, by triangulating signals across two independent databases (MHRA and WHO VigiAccess), strengthening confidence in the signals that are replicated; and third, by systematically integrating disproportionality analysis with comparative pharmacological, physicochemical and pharmacokinetic profiling to provide mechanistic plausibility for the observed generation-specific differences, while applying a pre-specified signal threshold that distinguishes confirmed signals from elevated but statistically weaker associations.

### Confirmed and expected ADR signals

#### Eye disorders - erlotinib

Erlotinib was consistently associated with eye disorders in both the Yellow Card (ROR 4.18, 95% CI 2.19–8.00) and VigiAccess (ROR 4.33, 95% CI 2.23–8.39) databases, the strongest signal detected for any drug–SOC combination in this study. At PT level, lid, lash and lacrimal structural disorders showed a particularly strong signal (ROR 7.84, 95% CI 1.44–42.78), although the wide confidence interval reflects the small number of reports (*n* = 12).

This signal is biologically plausible. EGFR is highly expressed in ocular tissues, including the corneal, limbal and conjunctival epithelium and the meibomian glands, where it regulates cell proliferation, migration, and growth [42]. Erlotinib inhibits WT EGFR with high potency (IC_50_ 2 nM) and achieves a steady-state C_max_ of approximately 5,337 nM, representing a C_max_:IC_50_ ratio of roughly 2,700-fold, the largest margin of any of the three drugs, such that at therapeutic concentrations erlotinib provides sustained, near-saturating, inhibition of EGFR in all tissues expressing the wild-type receptor, including the eye. Although erlotinib binding is reversible, allowing periodic receptor recovery between doses, the magnitude of target coverage is sufficient for ocular epithelial cells to experience near-continuous EGFR suppression during treatment. EGFR signalling also regulates the growth cycle of ocular hair follicles, and its inhibition may produce abnormal eyelash growth (trichomegaly) causing mechanical irritation of the ocular surface, consistent with the PT-level lid/lash/lacrimal signal [42].

By contrast, osimertinib’s WT EGFR IC_50_ (938 nM) is approximately 470-fold less potent than erlotinib’s, and its steady-state C_max_ (620–640 nM) falls below this threshold, such that osimertinib does not meaningfully inhibit WT EGFR in ocular tissue at therapeutic concentrations, a plausible pharmacological explanation for the absence of an eye disorder signal for osimertinib in either database. Afatinib, despite potent WT EGFR inhibition (IC50 31 nM), also did not generate an eye disorder signal; its C_max_ (94.7 nM) gives a C_max_:IC_50_ ratio of only approximately 3-fold, far below erlotinib’s margin, and its irreversible binding mechanism means that once target occupancy is achieved, further increases in plasma concentration do not proportionally increase target engagement. This pharmacokinetic–pharmacodynamic distinction may explain why afatinib’s recognised ocular toxicity did not reach the disproportionality threshold in either database in this analysis.

#### Gastrointestinal toxicity - afatinib

Gastrointestinal disorders showed elevated reporting for afatinib in both VigiAccess (ROR 1.62) and the Yellow Card scheme (ROR 1.63); although these SOC-level associations did not reach the pre-specified ROR ≥ 2 threshold, the PT-level finding for diarrhoea (Yellow Card ROR 2.79, 95% CI 1.74–4.46) met the signal criteria and is consistent with clinical trial data associating a higher incidence of gastrointestinal toxicity with second-generation EGFR-TKIs [15].

The pharmacological basis for afatinib’s gastrointestinal toxicity is multifactorial. First, afatinib irreversibly inhibits EGFR by covalent bonding to Cys797 in the kinase domain; once bound, receptor function cannot recover until new protein is synthesised, a process that outlasts the drug’s effective 37 h half-life. Second, afatinib is a pan-ErbB inhibitor with potent activity against ErbB4 (IC_50_ 2 nM) in addition to EGFR; because ErbB receptors are co-expressed in intestinal epithelium, simultaneous irreversible inhibition of multiple family members may compound disruption of epithelial homeostasis. Third, afatinib has the largest volume of distribution of the three drugs (2,370 L), indicating extensive tissue penetration, and the apparent terminal half-life of 344 h estimated after prolonged dosing reflects slow dissociation from covalently bound targets in tissue, which is likely to contribute to sustained gastrointestinal exposure. Mechanistically, afatinib-associated diarrhoea has been linked to excessive intestinal chloride secretion mediated by CFTR channels, crypt epithelial damage and alterations in intestinal microbiota [37], and EGFR inhibition more broadly has been associated with increased IgA secretion and altered expression of antimicrobial peptides such as α-defensin-5 [37].

The absence of a confirmed gastrointestinal signal for erlotinib and osimertinib is pharmacologically informative. Erlotinib, despite potent WT EGFR inhibition, binds reversibly, allowing GI epithelial cells to recover between doses. Osimertinib, while binding irreversibly, has approximately 30-fold lower potency against WT EGFR (IC_50_ 938 nM) than afatinib (IC_50_ 31 nM), and its C_max_ (620–640 nM) remains below this threshold. This pattern suggests that the combination of irreversible binding, broad ErbB family inhibition and sustained tissue exposure, rather than any single factor, underlies the relatively greater gastrointestinal reporting observed for afatinib.

#### Respiratory disorders, pneumonitis, and pulmonary embolism - osimertinib

Respiratory, thoracic and mediastinal disorders met the signal threshold for osimertinib in VigiAccess (ROR 2.04, 95% CI 1.96–2.12) and showed elevated but sub-threshold reporting in the Yellow Card scheme at SOC level (ROR 1.83); at PT level, pneumonitis met the signal threshold in the Yellow Card scheme (ROR 7.67, 95% CI 3.55–16.57), representing a recognised but uncommon ADR of osimertinib [43].

Pneumonitis encompasses a range of interstitial lung diseases, the precise mechanism of which remains incompletely understood [43]. Proposed explanations include immune-mediated facilitation of inflammatory signalling and disruption of EGFR-dependent epithelial repair in the alveolus, since EGFR signalling is important for lung tissue regeneration and its inhibition may impair this repair process, resulting in structural lung damage [44]. The detection of this signal specifically for osimertinib, rather than erlotinib or afatinib, may reflect several contributing factors rather than a single mechanism: osimertinib is increasingly used as first-line therapy, meaning patients are exposed for longer durations, which may increase cumulative risk; and its favourable BBB penetration (unbound brain:plasma Kp,_uu_ 0.21), reflecting weak P-glycoprotein/BCRP substrate status, may also facilitate greater tissue distribution more broadly, although this has not been directly demonstrated for lung tissue.

Pulmonary embolism was also identified at PT level for osimertinib (ROR 3.94, 95% CI 1.07– 14.52), meeting the pre-specified signal threshold, although the wide confidence interval and small number of reports (*n* = 6) warrant cautious interpretation. NSCLC itself is associated with an increased risk of thromboembolic events [53], so this signal may in part be disease-related rather than drug-related; however, an off-target contribution cannot be excluded, and this finding is regarded as hypothesis-generating.

#### Skin and vascular associations - erlotinib and afatinib

Skin and subcutaneous tissue disorders showed elevated reporting for erlotinib in both databases (Yellow Card ROR 1.56; VigiAccess ROR 1.66) and for afatinib in VigiAccess (ROR 1.16), although none of these SOC-level associations met the pre-specified ROR ≥ 2 threshold; erlotinib-associated rash at PT level (ROR 1.81) likewise fell short of the threshold despite its well-established clinical relevance. This illustrates an important general point: a well-recognised class-effect toxicity can still fail to reach a conservative disproportionality threshold, particularly when the comparator group (the other two EGFR-TKIs in this analysis) also causes the same ADR to some degree, reducing the relative disproportion.

EGFR, also known as ErbB1, is highly expressed in keratinocytes and regulates proliferation, differentiation, and migration [45]; its inhibition produces inflammatory skin reactions, typically beginning within days of treatment initiation, and has been widely reported for erlotinib [46]. Erlotinib’s skin toxicity may be further compounded by off-target inhibition of serine/threonine kinase 10 (STK10), which enhances lymphocytic activity and may contribute to the inflammatory component of the rash [55]; this off-target effect has not been reported for afatinib or osimertinib. Osimertinib’s WT-sparing selectivity (IC_50_ 938 nM, above its therapeutic C_max_) provides a plausible pharmacological explanation for the absence of a skin disorder signal, since at therapeutic concentrations osimertinib does not meaningfully inhibit EGFR in keratinocytes; rare but distinct dermatological reactions have nonetheless been reported with osimertinib in individual case reports [48].

Vascular disorders showed elevated reporting for erlotinib (ROR 1.38) and osimertinib (ROR 1.29) in VigiAccess, although neither met the pre-specified signal threshold. The mechanism underlying vascular toxicity with EGFR-TKIs is not fully established, but off-target inhibition of ErbB receptors expressed in vascular endothelium, and inhibitory activity against HER2 (osimertinib IC_50_ 4.80 nM, erlotinib IC_50_ 5.90 nM), a receptor involved in cardiomyocyte survival signalling [49,50], are plausible contributing factors; afatinib’s weaker HER2 coverage relative to its C_max_ may explain why it did not show a comparable association.

Immune system disorders showed elevated reporting for osimertinib in VigiAccess (ROR 1.90, 95% CI 1.56–2.31) but not in the Yellow Card scheme and did not meet the pre-specified signal threshold in either database. Immune system disorders are not commonly associated with osimertinib monotherapy; however, their incidence has been reported to increase when osimertinib is combined with immune checkpoint PD-(L)1 inhibitors, with EGFR and PD-(L)1 both expressed on macrophages and combined inhibition potentially shifting macrophage polarisation from a pro-inflammatory M1 towards an anti-inflammatory M2 phenotype [47]. This could plausibly impair immune surveillance in immunocompromised cancer patients, although causality cannot be inferred from spontaneous reporting data, and the absence of a comparable signal in the UK-specific Yellow Card data may reflect differences in combination-therapy prescribing practices between regions.

### Atypical and hypothesis-generating signals

#### Neoplasm-related terms and confounding by indication

Neoplasm-related terms showed elevated reporting for both afatinib and osimertinib and, for osimertinib, formally met the pre-specified statistical signal criteria at both SOC and PT level (Section 3.3.2). We do not interpret this as a genuine drug safety signal. EGFR-TKIs are not established to cause secondary malignancy; given the advanced disease stage of most NSCLC patients, disease progression, acquisition of resistance mutations, and natural disease biology are much more probable explanations [51]. The disproportionately stronger association for osimertinib most plausibly reflects its common use in later-line, *T790M*-resistant disease, where disease progression is intrinsically more frequent, rather than drug-related harm. Spontaneous reporting systems cannot distinguish disease progression from a drug-induced event, and this limitation is particularly acute for neoplasm-related MedDRA terms in an oncology population where every patient has, by definition, active malignancy. We therefore present these findings descriptively for transparency but explicitly exclude them from our interpretation of drug-specific ADR signals and recommend that future pharmacovigilance studies in oncology indications pre-specify neoplasm-related SOCs as a recognised source of confounding by indication, analogous to the exclusion of “Investigations” and “Surgical and medical procedures” applied in this study (**Table 1**).

#### Psychiatric disorders - afatinib

Psychiatric disorders were confirmed as a signal for afatinib in both the Yellow Card scheme (ROR 3.31, 95% CI 1.62–6.76) and VigiAccess (ROR 3.26, 95% CI 1.57–6.75), despite the absence of a clear mechanistic link between EGFR inhibition and mental health outcomes.

This finding is unexpected because afatinib has the poorest BBB penetration of the three drugs: it is a strong P-glycoprotein/BCRP substrate with a very low Kp,_uu_, making direct CNS pharmacological effects unlikely at therapeutic concentrations. Several alternative explanations merit consideration. First, these events may reflect cancer-related psychological distress, which is highly prevalent in advanced NSCLC and may be attributed to the drug in spontaneous reports. Second, concomitant medications commonly used in oncology (corticosteroids, opioids, and other psychoactive drugs) may contribute. Third, the modest number of reports (*n* = 12 in the Yellow Card scheme, *n* = 319 in VigiAccess) means the signal, while meeting the pre-specified statistical threshold in both databases which strengthens confidence that this is a genuine reporting pattern rather than a chance finding in one dataset, is nonetheless statistically fragile, as reflected in the relatively wide confidence intervals. Given the lack of pharmacological plausibility for a direct CNS drug effect, this signal should be considered hypothesis-generating rather than established and warrants targeted clinical and mechanistic follow-up.

By contrast, osimertinib, which has excellent BBB penetration confirmed by ¹¹C-PET imaging in healthy volunteers [52] and demonstrated clinical efficacy against leptomeningeal metastases [54], did not generate a psychiatric disorder signal in either database. This is reassuring but does not exclude the possibility of CNS effects at the individual patient level, and the clinical relevance of osimertinib’s CNS penetration in relation to psychiatric adverse events warrants further investigation.

#### Pulmonary embolism - osimertinib

As noted, pulmonary embolism met the pre-specified signal threshold for osimertinib at PT level (ROR 3.94, 95% CI 1.07–14.52), representing another atypical finding. Given the well-established disease-related risk of thromboembolism in NSCLC [53], an off-target drug contribution cannot be confirmed from spontaneous reporting data alone, and this signal, like the psychiatric disorder signal above, should be regarded as hypothesis-generating.

### Linking ADR signals to pharmacology, target selectivity, and structure

The generation-specific ADR patterns observed in this study can be understood through three interconnected pharmacological dimensions: target selectivity, binding mechanism, and pharmacokinetic behaviour.

#### Target selectivity and wild-type EGFR inhibition

Erlotinib, a first-generation reversible inhibitor, exhibits the highest potency against WT EGFR (IC_50_ 2 nM) and achieves by far the greatest Cmax:IC_50_ ratio of the three drugs (∼2,700-fold), providing a clear pharmacological explanation for its confirmed ocular signal and its elevated skin reporting. Afatinib, a second-generation irreversible pan-ErbB inhibitor, combines potent WT EGFR inhibition (IC_50_ 31 nM) with activity against ErbB2, ErbB3 and ErbB4, producing broader epithelial disruption manifesting as gastrointestinal toxicity. Osimertinib, designed to spare WT EGFR (IC50 938 nM, above its therapeutic C_max_), shows comparatively fewer confirmed epithelial toxicity signals, consistent with its improved tolerability profile in clinical trials [59].

#### Binding reversibility and tissue exposure

The distinction between reversible (erlotinib) and irreversible (afatinib, osimertinib) binding has implications for ADR persistence and severity. Erlotinib’s reversible binding allows target recovery between doses, potentially explaining why its toxicities, while frequent, are often manageable with dose modification. Afatinib’s irreversible pan-ErbB inhibition, combined with its large volume of distribution (2,370 L) and apparent terminal half-life of 344 h after prolonged dosing, creates sustained tissue-level target suppression that may explain the severity of its gastrointestinal toxicity. Osimertinib combines irreversible binding with mutant selectivity, a profile that reduces epithelial toxicity while potentially enabling atypical toxicities through off-target cysteine-reactive binding to non-EGFR kinases [58,60].

#### Structural features

Erlotinib and afatinib both contain a quinazoline core, associated with broader kinase inhibition, and potentially increased off-target effects [56]. Afatinib and osimertinib additionally contain a Michael acceptor (acrylamide warhead) enabling covalent binding to Cys797 in the EGFR kinase domain [57]; while this covalent mechanism is designed for target engagement, Michael acceptors may also react with other cysteine-containing kinases, potentially prolonging off-target inhibition [60]. Osimertinib’s pyrimidine core, distinct from the quinazoline scaffold of the other two drugs, combined with its dimethylamine side chains, confers mutant selectivity through steric complementarity with the *T790M* binding pocket while creating clashes with the WT EGFR conformation [9,17].

#### Metabolic considerations

Differences in CYP_450_ metabolism contribute further to inter-drug variability. Erlotinib’s metabolism by CYP1A2 means that smoking status significantly affects drug exposure, with smokers showing substantially lower AUC and C_max_ than non-smokers [14], potentially leading to subtherapeutic exposure during active smoking or supratherapeutic exposure following smoking cessation. Afatinib’s minimal hepatic metabolism and predominantly unchanged faecal elimination (85.4%) gives it low drug-drug interaction (DDI) potential but also limited capacity for dose adjustment via metabolic modulation. Osimertinib’s CYP3A4/CYP3A5-mediated metabolism generates two active metabolites (AZ5104 and AZ7550) that retain EGFR activity and may contribute to both efficacy and toxicity, a factor not captured by parent-compound pharmacokinetic parameters alone.

Taken together, these findings suggest that differences in ADR profiles between first-, second-and third-generation EGFR-TKIs reflect a combination of structural features, target selectivity, and pharmacokinetic behaviour. Broad inhibition of wild-type EGFR and ErbB family members, irreversible binding mechanisms, and sustained tissue exposure appear to contribute to epithelial toxicities, whereas enhanced central nervous system penetration and off-target kinase activity may be associated with atypical signals. While these associations provide mechanistic plausibility, they should be considered hypothesis-generating and interpreted in the context of the inherent limitations of spontaneous reporting pharmacovigilance.

### Limitations

This study has limitations typical of pharmacovigilance analyses based on spontaneous reporting. Under-reporting of ADRs is common, particularly for non-serious or well-established reactions, and reporting bias may differ between an older drug such as erlotinib (licensed 2005) and a newer drug such as osimertinib (licensed 2021), owing to recency bias, heightened clinical awareness and enhanced post-marketing surveillance for newly approved medicines. Declining reporting after the second year of marketing (the “Weber effect”) has been described, although reporting rates for oncology drugs are not always consistent with this pattern [61]. As previously noted, erlotinib is also licensed for pancreatic cancer, meaning that not all erlotinib ADRs captured in this analysis necessarily arose from NSCLC treatment. Disproportionality analysis using the ROR identifies statistical association rather than causality [30].

Both the MHRA Yellow Card scheme and WHO VigiAccess lack detailed patient-level information, including co-morbidities, cancer stage, treatment line, concomitant medications and confirmation of therapeutic indication; consequently, some reported ADRs may reflect disease progression rather than a drug-related effect, a limitation that is particularly relevant to the neoplasm-related findings discussed. WHO VigiAccess provides only aggregated data without access to individual case safety reports, does not stratify by seriousness, and reflects global variability in reporting practices. Delayed reporting and wide confidence intervals for rare events may result in both false-positive and false-negative signals. No correction for multiple statistical comparisons was applied, and no sensitivity analyses restricting to serious reports only, or excluding reports from the first year of marketing, were performed; both would be valuable additions in future studies. As a result, the present findings should be interpreted as hypothesis-generating and require confirmation through controlled observational or mechanistic studies; where the discussion links structural or pharmacokinetic features to specific ADR patterns, these associations are intended as plausible mechanistic hypotheses rather than established causal relationships.

Future studies should aim to validate the atypical ADR signals identified here using controlled observational designs with adjustment for drug exposure, treatment duration and patient-specific factors including age and sex. Integration of patient-level clinical data, mutation status and concomitant therapies would give further insight into the epidemiology of these associations and help distinguish drug-related effects from disease progression. Mechanistic studies exploring off-target kinase inhibition and tissue-specific drug exposure would also aid interpretation of the toxicities identified.

## Conclusions

This dual-database pharmacovigilance study demonstrates that EGFR-TKI toxicity profiles differ across generations used in NSCLC. Using a pre-specified, conservative signal threshold, confirmed generation-specific signals included eye disorders for erlotinib, psychiatric disorders and diarrhoea for afatinib, and pneumonitis and pulmonary embolism for osimertinib, while several well-recognised class-effect toxicities (gastrointestinal and dermatological reactions) showed elevated but sub-threshold reporting. These patterns align with known pharmacological characteristics, including target selectivity, binding reversibility, structural features, and pharmacokinetic behaviour.

These findings have potential clinical relevance, for example, differences in ADR profiles between EGFR-TKIs may inform treatment selection, patient counselling, and monitoring strategies in NSCLC, particularly in patients with pre-existing gastrointestinal, ocular, or dermatological conditions. Importantly, we explicitly distinguish genuine drug-related signals from neoplasm-related findings that reflect confounding by indication rather than a safety signal, a distinction we recommend for future pharmacovigilance research in oncology. While the results are hypothesis-generating and inherently limited by the nature of spontaneous reporting data, they provide a basis for future mechanistic and observational studies aimed at optimising EGFR-TKI safety and patient outcomes.

## Data Availability Statement

The data supporting the findings of this study are available within this article and Supplementary Information Tables A1-A4. The underlying datasets are available via the MHRA Yellow Card interactive Drug Analysis Profiles (iDAPs) repository (https://yellowcard.mhra.gov.uk/idaps) and the WHO VigiAccess database (https://www.vigiaccess.org).

## Author Contributions

O.A. carried out data acquisition, analysis, and interpretation, and drafted and revised the manuscript. A.M.J. conceived and designed the study, contributed to analysis and interpretation of data, and supervised, drafted, and revised the manuscript. All authors gave final approval of the version to be published and agree to be accountable for all aspects of the work, ensuring that questions related to the accuracy or integrity of any part of the work are appropriately investigated and resolved.

## Data Availability

https://www.vigiaccess.org

## Acknowledgements

We thank the electronic medicines compendium (emc), the European Medicines Agency (EMA), the US Food and Drug Administration (FDA), ChEMBL, the WHO Uppsala Monitoring Centre (VigiAccess) and the MHRA Yellow Card scheme for open-access public data that underpinned this research. We thank Mr Pono Pono, University of Birmingham, for statistical advice.

## Notes

### Competing Interest Statement

The authors have declared no competing interest.

### Author Declarations

The underlying datasets are available via the MHRA Yellow Card interactive Drug Analysis Profiles (iDAPs) repository (https://yellowcard.mhra.gov.uk/idaps) and the WHO VigiAccess database (https://www.vigiaccess.org).

