## Supplementary Information for "Pharmacovigilance and Mechanistic Analysis of Adverse Reaction Differences Across EGFR Tyrosine Kinase Inhibitor Generations in Non-Small Cell Lung Cancer"


Abbreviations: Erl: Erlotinib. Afa: Afatinib. Osi: Osimertinib. *n*: number of events. ROR: reporting odd ratios. CI: confidence interval. ROR: reporting odds ratio. Approx SE: approximate standard error. LnROR: natural log of reporting odds ratio. CI: confidence interval. DDD: daily drug doses. ADRs: adverse drug reactions. MHRA: medicines and healthcare products regulatory agency. WHO: world health organisation.

**Appendix A:** MHRA Yellow Card disproportionality analysis showing SOC level raw data.

***Table A1.*** *Disproportionality metrics at SOC level for adverse event reporting associated with EGFR-TKIs in MHRA Yellow Card data.*

| Drug | Event | A | B | C | D | ROR | Approx SE | LnROR | 95% CI lower | 95% CI upper | Signal? |
|---|---|---|---|---|---|---|---|---|---|---|---|
| Osi | Blood and lymphatic system disorders | 6 | 530 | 13 | 1116 | 1.01333473 | 0.480313333 | 0.013246605 | 0.395277382 | 2.597789102 | No Signal |
| Afa | Blood and lymphatic system disorders | 1 | 265 | 18 | 1381 | 0.421896473 | 0.851593242 | -0.86299532 | 0.079490475 | 2.239219649 | No Signal |
| Erl | Blood and lymphatic system disorders | 12 | 851 | 7 | 795 | 1.557056175 | 0.4645049 | 0.442796971 | 0.626483631 | 3.869891909 | No Signal |
| Osi | Cardiac disorders | 19 | 389 | 31 | 1098 | 1.745889113 | 0.294118699 | 0.557263946 | 0.980974607 | 3.10724536 | No Signal |
| Afa | Cardiac disorders | 6 | 212 | 44 | 1355 | 0.931738268 | 0.426335175 | -0.070703332 | 0.404008131 | 2.148808734 | No Signal |
| Erl | Cardiac disorders | 25 | 669 | 25 | 777 | 1.161314414 | 0.284975785 | 0.149552479 | 0.664314192 | 2.030140531 | No Signal |

| | | | | | | | | | | | |
|---|---|---|---|---|---|---|---|---|---|---|---|
| Osi | Eye disorders | 8 | 528 | 52 | 1077 | 0.330089652 | 0.373516934 | -1.108390989 | 0.158740549 | 0.686397894 | No Signal |
| Afa | Eye disorders | 3 | 263 | 57 | 1342 | 0.310122927 | 0.554658035 | -1.170786521 | 0.10456815 | 0.919746884 | No Signal |
| Erl | Eye disorders | 49 | 814 | 11 | 791 | 4.182800865 | 0.33113398 | 1.430981085 | 2.185748831 | 8.004498426 | Signal Detected |
| Osi | Gastrointestinal disorders | 58 | 478 | 191 | 938 | 0.599155324 | 0.159597442 | -0.512234409 | 0.438215099 | 0.81920295 | No Signal |
| Afa | Gastrointestinal disorders | 55 | 211 | 194 | 1205 | 1.626410691 | 0.169461116 | 0.486375557 | 1.166761275 | 2.267140497 | Signal Detected |
| Erl | Gastrointestinal disorders | 136 | 727 | 113 | 689 | 1.139824697 | 0.137700678 | 0.130874476 | 0.870211622 | 1.492970568 | No Signal |
| Osi | Infections and infestations | 17 | 519 | 74 | 1055 | 0.477259368 | 0.270994506 | -0.739695188 | 0.280594582 | 0.811763728 | No Signal |
| Afa | Infections and infestations | 18 | 248 | 73 | 1326 | 1.343585321 | 0.269142164 | 0.295341654 | 0.792805794 | 2.277003436 | No Signal |
| Erl | Infections and infestations | 56 | 807 | 35 | 767 | 1.512710941 | 0.220021478 | 0.413903366 | 0.98281072 | 2.328316474 | No Signal |
| Osi | Metabolism and nutrition disorders | 9 | 527 | 26 | 1103 | 0.749941876 | 0.381838905 | -0.287759574 | 0.354813199 | 1.585095533 | No Signal |
| Afa | Metabolism and nutrition disorders | 6 | 260 | 29 | 1370 | 1.159211425 | 0.438534937 | 0.147739968 | 0.490765673 | 2.738111489 | No Signal |
| Erl | Metabolism and nutrition disorders | 20 | 843 | 15 | 787 | 1.234774461 | 0.340223448 | 0.210888331 | 0.633845765 | 2.405424244 | No Signal |
| Osi | Musculoskeletal and connective tissue disorders | 13 | 523 | 19 | 1110 | 1.46859158 | 0.358003967 | 0.384303833 | 0.728051172 | 2.962375878 | No Signal |
| Afa | Musculoskeletal and connective tissue disorders | 0 | 266 | 32 | 1367 | 0.078943571 | 1.426622878 | -2.539021979 | 0.004818933 | 1.29325056 | No Signal |
| Erl | Musculoskeletal and connective tissue disorders | 19 | 844 | 13 | 789 | 1.350371686 | 0.357500885 | 0.300379878 | 0.670104334 | 2.721223543 | No Signal |
| Osi | Neoplasms benign, malignant and unspecified (incl cysts and polyps) | 73 | 463 | 50 | 1079 | 3.389759364 | 0.191026895 | 1.220758935 | 2.331113899 | 4.929175084 | Signal Detected |

| | | | | | | | | | | | |
|---|---|---|---|---|---|---|---|---|---|---|---|
| Afa | Neoplasms benign, malignant and unspecified (incl cysts and polyps) | 30 | 236 | 93 | 1306 | 1.802048592 | 0.220172219 | 0.588924124 | 1.170447993 | 2.77447537 | Signal Detected |
| Erl | Neoplasms benign, malignant and unspecified (incl cysts and polyps) | 20 | 843 | 103 | 699 | 0.164254071 | 0.247098063 | -1.806340834 | 0.101200353 | 0.266593932 | No Signal |
| Osi | Nervous system disorders | 24 | 512 | 85 | 1044 | 0.584002282 | 0.23541633 | -0.537850388 | 0.368149418 | 0.926413702 | No Signal |
| Afa | Nervous system disorders | 21 | 245 | 88 | 1311 | 1.297812604 | 0.250293564 | 0.260680235 | 0.794616913 | 2.119659837 | No Signal |
| Erl | Nervous system disorders | 64 | 799 | 45 | 757 | 1.343112797 | 0.200131986 | 0.294989903 | 0.907312098 | 1.988237554 | No Signal |
| Osi | Psychiatric disorders | 6 | 530 | 26 | 1103 | 0.510216421 | 0.440877789 | -0.672920288 | 0.215016447 | 1.210701783 | No Signal |
| Afa | Psychiatric disorders | 12 | 254 | 20 | 1379 | 3.305141598 | 0.365287094 | 1.195479316 | 1.615293399 | 6.762833916 | Signal Detected |
| Erl | Psychiatric disorders | 14 | 849 | 18 | 784 | 0.723812099 | 0.354219468 | -0.323223452 | 0.361499866 | 1.44925076 | No Signal |
| Osi | Renal and urinary disorders | 8 | 528 | 19 | 1110 | 0.915920724 | 0.414393235 | -0.087825464 | 0.406554941 | 2.06346225 | No Signal |
| Afa | Renal and urinary disorders | 3 | 263 | 24 | 1375 | 0.74573055 | 0.575371782 | -0.293390937 | 0.241443538 | 2.303288202 | No Signal |
| Erl | Renal and urinary disorders | 16 | 847 | 11 | 791 | 1.339976914 | 0.387306012 | 0.292652386 | 0.627214057 | 2.862719851 | No Signal |
| Osi | Respiratory, thoracic, and mediastinal disorders | 78 | 458 | 96 | 1033 | 1.833637509 | 0.162019007 | 0.606301704 | 1.334750589 | 2.518992345 | Signal Detected |
| Afa | Respiratory, thoracic, and mediastinal disorders | 25 | 241 | 149 | 1250 | 0.883213195 | 0.225488617 | -0.124188663 | 0.567708979 | 1.374058852 | No Signal |
| Erl | Respiratory, thoracic, and mediastinal disorders | 71 | 792 | 103 | 699 | 0.609753273 | 0.162293783 | -0.494700875 | 0.443615675 | 0.838110722 | No Signal |
| Osi | Skin and subcutaneous tissue disorders | 73 | 463 | 206 | 923 | 0.709176677 | 0.147269792 | -0.343650592 | 0.531368575 | 0.946483444 | No Signal |

| | | | | | | | | | | | |
|---|---|---|---|---|---|---|---|---|---|---|---|
| Afa | Skin and subcutaneous tissue disorders | 36 | 230 | 243 | 1156 | 0.752087908 | 0.191590994 | -0.284902063 | 0.51663409 | 1.094848816 | No Signal |
| Erl | Skin and subcutaneous tissue disorders | 170 | 693 | 109 | 693 | 1.557077626 | 0.13372152 | 0.442810747 | 1.198075559 | 2.02365428 | Signal Detected |
| Osi | Vascular disorders | 10 | 526 | 10 | 1119 | 2.126305793 | 0.439623454 | 0.754386104 | 0.898277834 | 5.03316029 | No Signal |
| Afa | Vascular disorders | 0 | 266 | 20 | 1379 | 0.126252688 | 1.432919305 | -2.069469917 | 0.007612286 | 2.093949342 | No Signal |
| Erl | Vascular disorders | 10 | 853 | 10 | 792 | 0.928529584 | 0.439214829 | -0.074153036 | 0.392580329 | 2.196154835 | No Signal |

**Appendix A2:** MHRA Yellow Card disproportionality analysis showing PT level raw data.

***Table A2.*** *Disproportionality metrics at PT level for adverse event reporting associated with EGFR-TKIs in MHRA Yellow Card data.*

| PT | A | B | C | D | ROR | Approx SE | LnROR | 95% CI lower | 95% CI upper | Signal? |
|---|---|---|---|---|---|---|---|---|---|---|
| Abnormal sensation in eye (2) | 2 | 53130 | 10 | 56094 | 0.251377897 | 0.703757589 | -1.380797908 | 0.063281466 | 0.998567992 | No Signal |
| Altered visual depth perception (1) | 1 | 53131 | 1 | 56103 | 1.055936685 | 1.154716406 | 0.054428226 | 0.109830619 | 10.15201677 | No Signal |
| Angle closure glaucoma (1) | 1 | 53131 | 0 | 56104 | 3.16786652 | 1.633004382 | 1.153058339 | 0.129040335 | 77.7693137 | No Signal |
| Anisocoria (1) | 1 | 53131 | 0 | 56104 | 3.16786652 | 1.633004382 | 1.153058339 | 0.129040335 | 77.7693137 | No Signal |
| Anterior chamber cell (1) | 1 | 53131 | 0 | 56104 | 3.16786652 | 1.633004382 | 1.153058339 | 0.129040335 | 77.7693137 | No Signal |
| Arcus lipoides (1) | 1 | 53131 | 0 | 56104 | 3.16786652 | 1.633004382 | 1.153058339 | 0.129040335 | 77.7693137 | No Signal |
| Asthenopia (3) | 3 | 53129 | 0 | 56104 | 7.391966798 | 1.511870011 | 2.000393843 | 0.381795032 | 143.1165116 | No Signal |

| | | | | | | | | | | |
|---|---|---|---|---|---|---|---|---|---|---|
| Blepharospasm (2) | 2 | 53130 | 17 | 56087 | 0.150807916 | 0.676150507 | -1.891748329 | 0.04007497 | 0.567512028 | No Signal |
| Blindness (34) | 34 | 53098 | 48 | 56056 | 0.750967396 | 0.222802011 | -0.286393042 | 0.485252947 | 1.162181565 | No Signal |
| Blindness transient (2) | 2 | 53130 | 3 | 56101 | 0.754227798 | 0.828100798 | -0.282060838 | 0.148802047 | 3.822928404 | No Signal |
| Blindness unilateral (17) | 17 | 53115 | 9 | 56095 | 1.945455014 | 0.40304177 | 0.66549589 | 0.882968299 | 4.28644518 | No Signal |
| Cataract (36) | 36 | 53096 | 83 | 56021 | 0.461206296 | 0.198519501 | -0.773909838 | 0.312544613 | 0.680578833 | No Signal |
| Cataract nuclear (2) | 2 | 53130 | 0 | 56104 | 5.279876907 | 1.549205166 | 1.663902784 | 0.253462514 | 109.9851008 | No Signal |
| Chorioretinal scar (1) | 1 | 53131 | 0 | 56104 | 3.16786652 | 1.633004382 | 1.153058339 | 0.129040335 | 77.7693137 | No Signal |
| Conjunctival disorder (4) | 4 | 53128 | 3 | 56101 | 1.357661142 | 0.712722355 | 0.305763471 | 0.335822563 | 5.488743104 | No Signal |
| Conjunctival haemorrhage (4) | 4 | 53128 | 3 | 56101 | 1.357661142 | 0.712722355 | 0.305763471 | 0.335822563 | 5.488743104 | No Signal |
| Conjunctival hyperaemia (9) | 9 | 53123 | 3 | 56101 | 2.866443288 | 0.625311197 | 1.053071989 | 0.841526347 | 9.763802587 | No Signal |
| Conjunctival irritation (2) | 2 | 53130 | 2 | 56102 | 1.055937738 | 0.894447677 | 0.054429224 | 0.182923321 | 6.095474859 | No Signal |
| Conjunctival oedema (2) | 2 | 53130 | 0 | 56104 | 5.279876907 | 1.549205166 | 1.663902784 | 0.253462514 | 109.9851008 | No Signal |
| Conjunctival pallor (1) | 1 | 53131 | 0 | 56104 | 3.16786652 | 1.633004382 | 1.153058339 | 0.129040335 | 77.7693137 | No Signal |
| Conjunctival ulcer (3) | 3 | 53129 | 0 | 56104 | 7.391966798 | 1.511870011 | 2.000393843 | 0.381795032 | 143.1165116 | No Signal |
| Conjunctivitis allergic (4) | 4 | 53128 | 2 | 56102 | 1.900759479 | 0.788833867 | 0.642253532 | 0.405002983 | 8.920641943 | No Signal |

| | | | | | | | | | | |
|---|---|---|---|---|---|---|---|---|---|---|
| Corneal defect (2) | 2 | 53130 | 0 | 56104 | 5.279876907 | 1.549205166 | 1.663902784 | 0.253462514 | 109.9851008 | No Signal |
| Corneal disorder (16) | 16 | 53116 | 8 | 56096 | 2.050082477 | 0.422243736 | 0.717880025 | 0.896087068 | 4.690211827 | No Signal |
| Corneal epitheliopathy (3) | 3 | 53129 | 1 | 56103 | 2.463945015 | 0.975918848 | 0.90176373 | 0.363841403 | 16.68591037 | No Signal |
| Corneal epithelium defect (3) | 3 | 53129 | 0 | 56104 | 7.391966798 | 1.511870011 | 2.000393843 | 0.381795032 | 143.1165116 | No Signal |
| Corneal erosion (2) | 2 | 53130 | 0 | 56104 | 5.279876907 | 1.549205166 | 1.663902784 | 0.253462514 | 109.9851008 | No Signal |
| Corneal exfoliation (3) | 3 | 53129 | 0 | 56104 | 7.391966798 | 1.511870011 | 2.000393843 | 0.381795032 | 143.1165116 | No Signal |
| Corneal infiltrates (1) | 1 | 53131 | 0 | 56104 | 3.16786652 | 1.633004382 | 1.153058339 | 0.129040335 | 77.7693137 | No Signal |
| Corneal irritation (1) | 1 | 53131 | 0 | 56104 | 3.16786652 | 1.633004382 | 1.153058339 | 0.129040335 | 77.7693137 | No Signal |
| Corneal lesion (2) | 2 | 53130 | 3 | 56101 | 0.754227798 | 0.828100798 | -0.282060838 | 0.148802047 | 3.822928404 | No Signal |
| Corneal neovascularisation (1) | 1 | 53131 | 0 | 56104 | 3.16786652 | 1.633004382 | 1.153058339 | 0.129040335 | 77.7693137 | No Signal |
| Corneal oedema (4) | 4 | 53128 | 0 | 56104 | 9.504136198 | 1.490724276 | 2.251727093 | 0.511661253 | 176.539858 | No Signal |
| Corneal scar (1) | 1 | 53131 | 0 | 56104 | 3.16786652 | 1.633004382 | 1.153058339 | 0.129040335 | 77.7693137 | No Signal |
| Corneal thinning (3) | 3 | 53129 | 0 | 56104 | 7.391966798 | 1.511870011 | 2.000393843 | 0.381795032 | 143.1165116 | No Signal |
| Corneal toxicity (1) | 1 | 53131 | 0 | 56104 | 3.16786652 | 1.633004382 | 1.153058339 | 0.129040335 | 77.7693137 | No Signal |
| Dacryostenosis acquired (6) | 6 | 53126 | 2 | 56102 | 2.745644829 | 0.744233029 | 1.010015958 | 0.638469879 | 11.80723754 | No Signal |
| Dark circles under eyes (1) | 1 | 53131 | 1 | 56103 | 1.055936685 | 1.154716406 | 0.054428226 | 0.109830619 | 10.15201677 | No Signal |

| | | | | | | | | | | |
|---|---|---|---|---|---|---|---|---|---|---|
| Deposit eye (2) | 2 | 53130 | 1 | 56103 | 1.7599276 | 1.0328133 | 0.565272672 | 0.232459004 | 13.32426407 | No Signal |
| Diplopia (26) | 26 | 53106 | 28 | 56076 | 0.981825332 | 0.269926344 | -0.018341856 | 0.578453284 | 1.666480269 | No Signal |
| Ectropion (6) | 6 | 53126 | 0 | 56104 | 13.72871354 | 1.467611257 | 2.619489519 | 0.773346139 | 243.7169672 | No Signal |
| Eczema eyelids (3) | 3 | 53129 | 1 | 56103 | 2.463945015 | 0.975918848 | 0.90176373 | 0.363841403 | 16.68591037 | No Signal |
| Entropion (3) | 3 | 53129 | 1 | 56103 | 2.463945015 | 0.975918848 | 0.90176373 | 0.363841403 | 16.68591037 | No Signal |
| Episcleritis (2) | 2 | 53130 | 1 | 56103 | 1.7599276 | 1.0328133 | 0.565272672 | 0.232459004 | 13.32426407 | No Signal |
| Evelid bleeding (1) | 1 | 53131 | 1 | 56103 | 1.055936685 | 1.154716406 | 0.054428226 | 0.109830619 | 10.15201677 | No Signal |
| Exophthalmos (2) | 2 | 53130 | 1 | 56103 | 1.7599276 | 1.0328133 | 0.565272672 | 0.232459004 | 13.32426407 | No Signal |
| Extraocular muscle disorder (1) | 1 | 53131 | 0 | 56104 | 3.16786652 | 1.633004382 | 1.153058339 | 0.129040335 | 77.7693137 | No Signal |
| Eye allergy (4) | 4 | 53128 | 1 | 56103 | 3.167988932 | 0.942828476 | 1.15309698 | 0.499150896 | 20.1064527 | No Signal |
| Eye haemorrhage (19) | 19 | 53113 | 16 | 56088 | 1.24801433 | 0.334551593 | 0.221553752 | 0.647803823 | 2.404338646 | No Signal |
| Eye inflammation (18) | 18 | 53114 | 17 | 56087 | 1.116314754 | 0.333516968 | 0.110032862 | 0.580619063 | 2.146258553 | No Signal |
| Eye movement disorder (1) | 1 | 53131 | 0 | 56104 | 3.16786652 | 1.633004382 | 1.153058339 | 0.129040335 | 77.7693137 | No Signal |
| Eye oedema (1) | 1 | 53131 | 3 | 56101 | 0.452528161 | 0.975918848 | -0.792905283 | 0.066823115 | 3.064534432 | No Signal |
| Eye ulcer (4) | 4 | 53128 | 2 | 56102 | 1.900759479 | 0.788833867 | 0.642253532 | 0.405002983 | 8.920641943 | No Signal |
| Eyelash changes (2) | 2 | 53130 | 0 | 56104 | 5.279876907 | 1.549205166 | 1.663902784 | 0.253462514 | 109.9851008 | No Signal |

| | | | | | | | | | | |
|---|---|---|---|---|---|---|---|---|---|---|
| Eyelash hyperpigmentation (3) | 3 | 53129 | 0 | 56104 | 7.391966798 | 1.511870011 | 2.000393843 | 0.381795032 | 143.1165116 | No Signal |
| Eyelid disorder (9) | 9 | 53123 | 5 | 56099 | 1.824035245 | 0.535833919 | 0.601051215 | 0.638149136 | 5.21367873 | No Signal |
| Eyelid irritation (8) | 8 | 53124 | 2 | 56102 | 3.590593794 | 0.719502402 | 1.278317591 | 0.876422395 | 14.7102172 | No Signal |
| Eyelid pain (3) | 3 | 53129 | 0 | 56104 | 7.391966798 | 1.511870011 | 2.000393843 | 0.381795032 | 143.1165116 | No Signal |
| Eyelid ptosis (13) | 13 | 53119 | 6 | 56098 | 2.193399208 | 0.4774483 | 0.78545249 | 0.86041007 | 5.591519966 | No Signal |
| Eyelid rash (3) | 3 | 53129 | 4 | 56100 | 0.821271087 | 0.712722355 | -0.196902033 | 0.203144476 | 3.320229087 | No Signal |
| Eyelid retraction (1) | 1 | 53131 | 0 | 56104 | 3.16786652 | 1.633004382 | 1.153058339 | 0.129040335 | 77.7693137 | No Signal |
| Eyelid skin dryness (3) | 3 | 53129 | 1 | 56103 | 2.463945015 | 0.975918848 | 0.90176373 | 0.363841403 | 16.68591037 | No Signal |
| Eyelids pruritus (19) | 19 | 53113 | 19 | 56085 | 1.055955642 | 0.320313534 | 0.054446179 | 0.563623757 | 1.978345137 | No Signal |
| Foreign body sensation in eyes (9) | 9 | 53123 | 6 | 56098 | 1.543386926 | 0.509063809 | 0.433979304 | 0.569050691 | 4.18599475 | No Signal |
| Giant papillary conjunctivitis (1) | 1 | 53131 | 0 | 56104 | 3.16786652 | 1.633004382 | 1.153058339 | 0.129040335 | 77.7693137 | No Signal |
| Glare (3) | 3 | 53129 | 1 | 56103 | 2.463945015 | 0.975918848 | 0.90176373 | 0.363841403 | 16.68591037 | No Signal |
| Glaucoma (9) | 9 | 53123 | 13 | 56091 | 0.743019498 | 0.423525541 | -0.297032992 | 0.323957465 | 1.704168091 | No Signal |
| Halo vision (2) | 2 | 53130 | 1 | 56103 | 1.7599276 | 1.0328133 | 0.565272672 | 0.232459004 | 13.32426407 | No Signal |
| Iridocele (1) | 1 | 53131 | 0 | 56104 | 3.16786652 | 1.633004382 | 1.153058339 | 0.129040335 | 77.7693137 | No Signal |
| Iris adhesions (2) | 2 | 53130 | 0 | 56104 | 5.279876907 | 1.549205166 | 1.663902784 | 0.253462514 | 109.9851008 | No Signal |

| | | | | | | | | | | |
|---|---|---|---|---|---|---|---|---|---|---|
| Iris disorder (1) | 1 | 53131 | 0 | 56104 | 3.16786652 | 1.633004382 | 1.153058339 | 0.129040335 | 77.7693137 | No Signal |
| Keratitis (45) | 45 | 53087 | 44 | 56060 | 1.079732263 | 0.210918484 | 0.076713106 | 0.714132241 | 1.632501227 | No Signal |
| Keratitis interstitial (1) | 1 | 53131 | 0 | 56104 | 3.16786652 | 1.633004382 | 1.153058339 | 0.129040335 | 77.7693137 | No Signal |
| Keratopathy (2) | 2 | 53130 | 2 | 56102 | 1.055937738 | 0.894447677 | 0.054429224 | 0.182923321 | 6.095474859 | No Signal |
| Lacrimal disorder (2) | 2 | 53130 | 2 | 56102 | 1.055937738 | 0.894447677 | 0.054429224 | 0.182923321 | 6.095474859 | No Signal |
| Lacrimal haemorrhage (2) | 2 | 53130 | 0 | 56104 | 5.279876907 | 1.549205166 | 1.663902784 | 0.253462514 | 109.9851008 | No Signal |
| Lacrimation decreased (3) | 3 | 53129 | 0 | 56104 | 7.391966798 | 1.511870011 | 2.000393843 | 0.381795032 | 143.1165116 | No Signal |
| Lacrimation disorder (3) | 3 | 53129 | 1 | 56103 | 2.463945015 | 0.975918848 | 0.90176373 | 0.363841403 | 16.68591037 | No Signal |
| Lens disorder (1) | 1 | 53131 | 0 | 56104 | 3.16786652 | 1.633004382 | 1.153058339 | 0.129040335 | 77.7693137 | No Signal |
| Limbal stem cell deficiency (1) | 1 | 53131 | 0 | 56104 | 3.16786652 | 1.633004382 | 1.153058339 | 0.129040335 | 77.7693137 | No Signal |
| Macular degeneration (8) | 8 | 53124 | 15 | 56089 | 0.578993836 | 0.426848732 | -0.546463447 | 0.250803137 | 1.336641425 | No Signal |
| Macular hole (1) | 1 | 53131 | 1 | 56103 | 1.055936685 | 1.154716406 | 0.054428226 | 0.109830619 | 10.15201677 | No Signal |
| Maculopathy (1) | 1 | 53131 | 2 | 56102 | 0.633550719 | 1.0328133 | -0.456415222 | 0.083682175 | 4.796559289 | No Signal |
| Metamorphopsia (2) | 2 | 53130 | 3 | 56101 | 0.754227798 | 0.828100798 | -0.282060838 | 0.148802047 | 3.822928404 | No Signal |
| Mydriasis (2) | 2 | 53130 | 1 | 56103 | 1.7599276 | 1.0328133 | 0.565272672 | 0.232459004 | 13.32426407 | No Signal |

| | | | | | | | | | | |
|---|---|---|---|---|---|---|---|---|---|---|
| Night blindness (3) | 3 | 53129 | 0 | 56104 | 7.391966798 | 1.511870011 | 2.000393843 | 0.381795032 | 143.1165116 | No Signal |
| Ocular discomfort (8) | 8 | 53124 | 20 | 56084 | 0.437736803 | 0.408000246 | -0.826137456 | 0.19675068 | 0.973889945 | No Signal |
| Ocular ischaemic syndrome (1) | 1 | 53131 | 0 | 56104 | 3.16786652 | 1.633004382 | 1.153058339 | 0.129040335 | 77.7693137 | No Signal |
| Ocular toxicity (3) | 3 | 53129 | 9 | 56095 | 0.388988475 | 0.625311196 | -0.944205564 | 0.114198684 | 1.324989296 | No Signal |
| Ocular vascular disorder (3) | 3 | 53129 | 1 | 56103 | 2.463945015 | 0.975918848 | 0.90176373 | 0.363841403 | 16.68591037 | No Signal |
| Optic atrophy (2) | 2 | 53130 | 1 | 56103 | 1.7599276 | 1.0328133 | 0.565272672 | 0.232459004 | 13.32426407 | No Signal |
| Optic ischaemic neuropathy (3) | 3 | 53129 | 1 | 56103 | 2.463945015 | 0.975918848 | 0.90176373 | 0.363841403 | 16.68591037 | No Signal |
| Optic nerve disorder (1) | 1 | 53131 | 1 | 56103 | 1.055936685 | 1.154716406 | 0.054428226 | 0.109830619 | 10.15201677 | No Signal |
| Optic nerve infarction (1) | 1 | 53131 | 0 | 56104 | 3.16786652 | 1.633004382 | 1.153058339 | 0.129040335 | 77.7693137 | No Signal |
| Optic neuropathy (5) | 5 | 53127 | 1 | 56103 | 3.872059354 | 0.921152265 | 1.353786498 | 0.636562966 | 23.55280538 | No Signal |
| Orbital oedema (1) | 1 | 53131 | 7 | 56097 | 0.211164752 | 0.894447677 | -1.555116637 | 0.036580715 | 1.218963382 | No Signal |
| Panophthalmitis (1) | 1 | 53131 | 0 | 56104 | 3.16786652 | 1.633004382 | 1.153058339 | 0.129040335 | 77.7693137 | No Signal |
| Papilloedema (5) | 5 | 53127 | 4 | 56100 | 1.290617435 | 0.635670553 | 0.255120736 | 0.371281944 | 4.486330107 | No Signal |
| Parophthalmia (1) | 1 | 53131 | 0 | 56104 | 3.16786652 | 1.633004382 | 1.153058339 | 0.129040335 | 77.7693137 | No Signal |
| Periorbital oedema (8) | 8 | 53124 | 8 | 56096 | 1.055944056 | 0.485109027 | 0.054435207 | 0.408044727 | 2.732587326 | No Signal |

| | | | | | | | | | | |
|---|---|---|---|---|---|---|---|---|---|---|
| Photophobia (18) | 18 | 53114 | 27 | 56077 | 0.71025546 | 0.300756297 | -0.342130571 | 0.393917843 | 1.280629522 | No Signal |
| Photopsia (4) | 4 | 53128 | 4 | 56100 | 1.055939844 | 0.666694152 | 0.054431218 | 0.285849646 | 3.900683347 | No Signal |
| Pterygium (1) | 1 | 53131 | 0 | 56104 | 3.16786652 | 1.633004382 | 1.153058339 | 0.129040335 | 77.7693137 | No Signal |
| Punctate keratitis (2) | 2 | 53130 | 1 | 56103 | 1.7599276 | 1.0328133 | 0.565272672 | 0.232459004 | 13.32426407 | No Signal |
| Pupillary reflex impaired (1) | 1 | 53131 | 0 | 56104 | 3.16786652 | 1.633004382 | 1.153058339 | 0.129040335 | 77.7693137 | No Signal |
| Retinal degeneration (1) | 1 | 53131 | 0 | 56104 | 3.16786652 | 1.633004382 | 1.153058339 | 0.129040335 | 77.7693137 | No Signal |
| Retinal detachment (3) | 3 | 53129 | 18 | 56086 | 0.19971879 | 0.582927947 | -1.610844951 | 0.063711912 | 0.62606181 | No Signal |
| Retinal disorder (1) | 1 | 53131 | 1 | 56103 | 1.055936685 | 1.154716406 | 0.054428226 | 0.109830619 | 10.15201677 | No Signal |
| Retinal fibrosis (1) | 1 | 53131 | 0 | 56104 | 3.16786652 | 1.633004382 | 1.153058339 | 0.129040335 | 77.7693137 | No Signal |
| Retinal haemorrhage (6) | 6 | 53126 | 2 | 56102 | 2.745644829 | 0.744233029 | 1.010015958 | 0.638469879 | 11.80723754 | No Signal |
| Retinal oedema (1) | 1 | 53131 | 0 | 56104 | 3.16786652 | 1.633004382 | 1.153058339 | 0.129040335 | 77.7693137 | No Signal |
| Retinal tear (2) | 2 | 53130 | 6 | 56098 | 0.406100943 | 0.744233029 | -0.901153522 | 0.094434363 | 1.746376752 | No Signal |
| Retinal vein occlusion (2) | 2 | 53130 | 2 | 56102 | 1.055937738 | 0.894447677 | 0.054429224 | 0.182923321 | 6.095474859 | No Signal |
| Retinopathy (4) | 4 | 53128 | 1 | 56103 | 3.167988932 | 0.942828476 | 1.15309698 | 0.499150896 | 20.1064527 | No Signal |
| Scleral discolouration (2) | 2 | 53130 | 1 | 56103 | 1.7599276 | 1.0328133 | 0.565272672 | 0.232459004 | 13.32426407 | No Signal |

| | | | | | | | | | | |
|---|---|---|---|---|---|---|---|---|---|---|
| Scleral haemorrhage (1) | 1 | 53131 | 0 | 56104 | 3.16786652 | 1.633004382 | 1.153058339 | 0.129040335 | 77.7693137 | No Signal |
| Scleral hyperaemia (1) | 1 | 53131 | 0 | 56104 | 3.16786652 | 1.633004382 | 1.153058339 | 0.129040335 | 77.7693137 | No Signal |
| Scleritis (1) | 1 | 53131 | 1 | 56103 | 1.055936685 | 1.154716406 | 0.054428226 | 0.109830619 | 10.15201677 | No Signal |
| Serous retinal detachment (1) | 1 | 53131 | 1 | 56103 | 1.055936685 | 1.154716406 | 0.054428226 | 0.109830619 | 10.15201677 | No Signal |
| Strabismus (2) | 2 | 53130 | 1 | 56103 | 1.7599276 | 1.0328133 | 0.565272672 | 0.232459004 | 13.32426407 | No Signal |
| Swelling of eyelid (4) | 4 | 53128 | 3 | 56101 | 1.357661142 | 0.712722355 | 0.305763471 | 0.335822563 | 5.488743104 | No Signal |
| Vision blurred (188) | 188 | 52944 | 168 | 55936 | 1.181914027 | 0.106190974 | 0.167135181 | 0.959829966 | 1.455383573 | No Signal |
| Visual brightness (1) | 1 | 53131 | 0 | 56104 | 3.16786652 | 1.633004382 | 1.153058339 | 0.129040335 | 77.7693137 | No Signal |
| Visual field defect (6) | 6 | 53126 | 5 | 56099 | 1.247953641 | 0.579397087 | 0.221505122 | 0.400871975 | 3.885001665 | No Signal |
| Vitreous detachment (1) | 1 | 53131 | 0 | 56104 | 3.16786652 | 1.633004382 | 1.153058339 | 0.129040335 | 77.7693137 | No Signal |
| Vitreous disorder (1) | 1 | 53131 | 0 | 56104 | 3.16786652 | 1.633004382 | 1.153058339 | 0.129040335 | 77.7693137 | No Signal |
| Vitreous haemorrhage (2) | 2 | 53130 | 4 | 56100 | 0.586611164 | 0.788833867 | -0.533393091 | 0.124991759 | 2.75308276 | No Signal |
| Xanthopsia (1) | 1 | 53131 | 1 | 56103 | 1.055936685 | 1.154716406 | 0.054428226 | 0.109830619 | 10.15201677 | No Signal |
| Xerophthalmia (5) | 5 | 53127 | 8 | 56096 | 0.683219336 | 0.547267658 | -0.380939334 | 0.233731106 | 1.997118269 | No Signal |

**Appendix A3:** WHO VigiAccess disproportionality analysis showing SOC level

**Table A3.** Disproportionality metrics at SOC level for adverse event reporting associated with EGFR-TKIs in WHO VigiAccess.

| Drug | Event | A | B | C | D | ROR | Approx SE | LnROR | 95% CI lower | 95% CI upper | Signal? |
|---|---|---|---|---|---|---|---|---|---|---|---|
| Osi | Blood and lymphatic system disorders | 1334 | 30234 | 1648 | 76020 | 0.97184325 | 0.496359 | -0.028560752 | 0.367355566 | 2.571022173 | No Signal |
| Afa | Blood and lymphatic system disorders | 274 | 24262 | 2708 | 81992 | 0.28951782 | 1.029589 | -1.23953843 | 0.038483233 | 2.178106162 | No Signal |
| Erl | Blood and lymphatic system disorders | 1374 | 51758 | 1608 | 54496 | 1.60147725 | 0.478146 | 0.470926487 | 0.627357126 | 4.08814898 | No Signal |
| Osi | Cardiac disorders | 1454 | 30114 | 1275 | 76393 | 1.7299942 | 0.297273 | 0.548118053 | 0.966053054 | 3.098049226 | No Signal |
| Afa | Cardiac disorders | 215 | 24321 | 2514 | 82186 | 0.87156947 | 0.441417 | -0.137459706 | 0.366910644 | 2.07034969 | No Signal |
| Erl | Cardiac disorders | 1060 | 52072 | 1669 | 54435 | 1.16143498 | 0.287718 | 0.14965629 | 0.660821576 | 2.041294134 | No Signal |
| Osi | Eye disorders | 681 | 30887 | 2124 | 75544 | 0.31381119 | 0.383475 | -1.158963783 | 0.147995245 | 0.665409638 | No Signal |
| Afa | Eye disorders | 426 | 24110 | 2379 | 82321 | 0.26856114 | 0.596175 | -1.314 | 0.083477371 | 0.864007618 | No Signal |

| | | | | | | | | | | | |
|---|---|---|---|---|---|---|---|---|---|---|---|
| | | | | | | | | 676694 | | | |
| Erl | Eye disorders | 1698 | 51434 | 1107 | 54997 | 4.33 | 0.34 | 1.47 | 2.23 | 8.39 | Signal Detected |
| Osi | Gastrointestinal disorders | 4572 | 26996 | 17398 | 60270 | 0.59589476 | 0.16011 | -0.517691201 | 0.435392961 | 0.815563409 | No Signal |
| Afa | Gastrointestinal disorders | 7340 | 17196 | 14630 | 70070 | 1.62 | 0.17 | 0.48 | 1.16 | 2.26 | Signal Detected |
| Erl | Gastrointestinal disorders | 10058 | 43074 | 11912 | 44192 | 1.14063128 | 0.137947 | 0.131581861 | 0.870406974 | 1.494748719 | No Signal |
| Osi | Infections and infestations | 2340 | 29228 | 7057 | 70611 | 0.46698433 | 0.274248 | -0.761459586 | 0.272808638 | 0.799367506 | No Signal |
| Afa | Infections and infestations | 2799 | 21737 | 6598 | 78102 | 1.31838268 | 0.272104 | 0.276405741 | 0.773431543 | 2.247300231 | No Signal |
| Erl | Infections and infestations | 4258 | 48874 | 5139 | 50965 | 1.52069393 | 0.221295 | 0.419166762 | 0.985534069 | 2.34645366 | No Signal |
| Osi | Metabolism and nutrition disorders | 1533 | 30035 | 4337 | 73331 | 0.72449277 | 0.390355 | -0.32228349 | 0.337098966 | 1.557079179 | No Signal |
| Afa | Metabolism and nutrition disorders | 1447 | 23089 | 4423 | 80277 | 1.09018568 | 0.45357 | 0.086348027 | 0.44814042 | 2.652081255 | No Signal |
| Erl | Metabolism and nutrition disorders | 2890 | 50242 | 2980 | 53124 | 1.24476078 | 0.345143 | 0.218943363 | 0.632840711 | 2.448371859 | No Signal |

| | | | | | | | | | | | |
|---|---|---|---|---|---|---|---|---|---|---|---|
| Osi | Musculoskeletal and connective tissue disorders | 1507 | 30061 | 2163 | 75505 | 1.45214854 | 0.363824 | 0.373044209 | 0.711734719 | 2.962810881 | No Signal |
| Afa | Musculoskeletal and connective tissue disorders | 505 | 24031 | 3165 | 81535 | 0 | - | - | - | - | No Signal |
| Erl | Musculoskeletal and connective tissue disorders | 1658 | 51474 | 2012 | 54092 | 1.36629603 | 0.363328 | 0.312103448 | 0.670307596 | 2.784937604 | No Signal |
| Osi | Neoplasms benign, malignant and unspecified (incl cysts and polyps) | 3589 | 27979 | 5894 | 71774 | 3.40 | 0.19 | 1.22 | 2.34 | 4.96 | Signal Detected |
| Afa | Neoplasms benign, malignant and unspecified (incl cysts and polyps) | 1875 | 22661 | 7608 | 77092 | 1.79 | 0.22 | 0.58 | 1.16 | 2.76 | Signal Detected |
| Erl | Neoplasms benign, malignant and unspecified (incl cysts and polyps) | 4019 | 49113 | 5464 | 50640 | 0.16100612 | 0.249651 | -1.82631293 | 0.098704102 | 0.262633151 | No Signal |
| Osi | Nervous system disorders | 2443 | 29125 | 4227 | 73441 | 0.57573529 | 0.237365 | -0.552107283 | 0.361554118 | 0.916795335 | No Signal |
| Afa | Nervous system disorders | 992 | 23544 | 5678 | 79022 | 1.27694805 | 0.25264 | 0.244472896 | 0.77825427 | 2.095197405 | No Signal |
| Erl | Nervous system disorders | 3235 | 49897 | 3435 | 52669 | 1.34746211 | 0.201047 | 0.298222901 | 0.908619646 | 1.998255414 | No Signal |
| Osi | Psychiatric disorders | 682 | 30886 | 1472 | 76196 | 0.48026125 | 0.455984 | -0.733425056 | 0.196487956 | 1.173867708 | No Signal |
| Afa | Psychiatric disorders | 319 | 24217 | 1835 | 82865 | 3.26 | 0.37 | 1.18 | 1.57 | 6.75 | Signal Detected |

| | | | | | | | | | | | |
|---|---|---|---|---|---|---|---|---|---|---|---|
| Erl | Psychiatric disorders | 1153 | 51979 | 1001 | 55103 | 0.7182306 | 0.359774 | -0.330964594 | 0.354828013 | 1.453817548 | No Signal |
| Osi | Renal and urinary disorders | 548 | 31020 | 1377 | 76291 | 0.978764327 | 0.050954675 | -0.021464394 | 0.885736695 | 1.081562516 | No Signal |
| Afa | Renal and urinary disorders | 460 | 24076 | 1465 | 83235 | 1.085530065 | 0.053944937 | 0.082068407 | 0.97661412 | 1.206592756 | No Signal |
| Erl | Renal and urinary disorders | 917 | 52215 | 1008 | 55096 | 0.959916797 | 0.04604213 | -0.040908668 | 0.877085111 | 1.050571085 | No Signal |
| Osi | Respiratory, thoracic, and mediastinal disorders | 5224 | 26344 | 6888 | 70780 | 2.04 | 0.02 | 0.71 | 1.96 | 2.12 | Signal Detected |
| Afa | Respiratory, thoracic, and mediastinal disorders | 1917 | 22619 | 10195 | 74505 | 0.619365344 | 0.026026299 | -0.479059964 | 0.58856288 | 0.651779856 | No Signal |
| Erl | Respiratory, thoracic, and mediastinal disorders | 4971 | 48161 | 7141 | 48963 | 0.707713131 | 0.019554806 | -0.34571645 | 0.681101547 | 0.735364467 | No Signal |
| Osi | Skin and subcutaneous tissue disorders | 3802 | 27766 | 18776 | 58892 | 0.429488975 | 0.019216515 | -0.845159207 | 0.413613392 | 0.445973905 | No Signal |
| Afa | Skin and subcutaneous tissue disorders | 5535 | 19001 | 17043 | 67657 | 1.16 | 0.02 | 0.15 | 1.12 | 1.20 | Signal Detected |

| | | | | | | | | | | | |
|---|---|---|---|---|---|---|---|---|---|---|---|
| Erl | Skin and subcutaneous tissue disorders | 13241 | 39891 | 9337 | 46767 | 1.66 | 0.02 | 0.51 | 1.61 | 1.71 | Signal Detected |
| Osi | Vascular disorders | 887 | 30681 | 1702 | 75966 | 1.29 | 0.04 | 0.25 | 1.19 | 1.40 | Signal Detected |
| Afa | Vascular disorders | 240 | 24296 | 2349 | 82351 | 0.34630784 | 0.068159239 | -1.060427189 | 0.303000962 | 0.395804419 | No Signal |
| Erl | Vascular disorders | 1462 | 51670 | 1127 | 54977 | 1.38 | 0.04 | 0.32 | 1.28 | 1.49 | Signal Detected |
| Osi | Immune system disorders | 176 | 31392 | 229 | 77439 | 1.90 | 0.10 | 0.64 | 1.56 | 2.31 | Signal Detected |
| Afa | Immune system disorders | 40 | 24496 | 365 | 84335 | 0.377293917 | 0.166710547 | -0.974730776 | 0.272127806 | 0.523102367 | No Signal |
| Erl | Immune system disorders | 189 | 52943 | 216 | 55888 | 0.923672629 | 0.099786853 | -0.079397568 | 0.759587467 | 1.123203269 | No Signal |

**Appendix A4:** Chi-squared analysis of adverse event reporting per 100,000 DDDs

| | Erlotinib | Afatinib | Osimertinib | Chi-value | Statistically significant |
|---|---|---|---|---|---|
| DDD | 139139 | 387801 | 2865611 | | |
| Total ADRs | 37 (26.59) | 71 (18.31) | 352 (12.28) | < 0.001 | Yes |
| Fatalities | 1 (0.72) | 9 (2.32) | 21 (0.73) | < 0.001 | Yes |
| Blood and lymphatic system disorders | 0 | 0 | 5 (0.17) | - | |
| Cardiac disorders | 0 | 2 (0.52) | 9 (0.31) | - | |
| Congenital, familial, and genetic disorders | 2 (1.44) | 0 | 6 (0.21) | - | |
| Ear and labyrinth disorders | 0 | 0 | 3 (0.10) | - | |
| Endocrine disorders | 0 | 0 | 1 (0.03) | - | |
| Eye disorders | 6 (4.31) | 0 | 7 (0.24) | <0.001 | Yes |
| Gastrointestinal disorders | 3 (2.16) | 9 (2.32) | 49 (1.71) | <0.001 | Yes |
| Hepatobiliary disorders | 0 | 3 (0.77) | 2 (0.07) | - | |
| Immune system disorders | 0 | 0 | 2 (0.07) | - | |
| Infections and infestations | 6 (4.31) | 2 (0.52) | 16 (0.56) | < 0.001 | Yes |
| Metabolism and nutrition disorders | 1 (0.72) | 0 | 5 (0.17) | - | |
| Musculoskeletal and connective tissue disorders | 0 | 0 | 12 (0.42) | - | |

| | | | | | |
|---|---|---|---|---|---|
| Neoplasms benign, malignant, and unspecified (incl cysts and polyps) | 5 (3.60) | 4 (1.03) | 66 (2.30) | < 0.001 | Yes |
| Nervous system disorders | 3 (2.16) | 0 | 14 (0.49) | - | |
| Psychiatric disorders | 0 | 0 | 5 (0.17) | - | |
| Renal and urinary disorders | 0 | 0 | 5 (0.17) | - | |
| Reproductive system and breast disorders | 0 | 0 | 5 (0.17) | - | |
| Respiratory, thoracic, and mediastinal disorders | 0 | 1 (0.26) | 64 (2.23) | - | |
| Skin and subcutaneous tissue disorders | 11 (7.91) | 9 (2.32) | 68 (2.37) | < 0.001 | Yes |
| Vascular disorders | 0 | 41 (10.57) | 8 (0.28) | < 0.001 | Yes |
